# OpenSpindleNet: An Open-Source Deep Learning Network for Reliable Sleep Spindle Detection

**DOI:** 10.1101/2025.04.13.25325696

**Authors:** Michal Sejak, Filip Mivalt, Vladimir Sladky, Vit Vsiansky, Diego Z. Carvalho, Erik K. St. Louis, Gregory A. Worrell, Vaclav Kremen

## Abstract

Sleep spindles, an oscillatory brain activity occurring during light non-rapid eye movement (NREM) sleep, are important for memory consolidation and cognitive functions. Accurate detection is important for understanding the role of spindles in sleep state physiology and brain health and for better understanding sleep and neurological disorders. However, manual spindle labeling of electroencephalography (EEG) data is time-consuming and impractical for most clinical and research settings and intracranial EEG (iEEG) presents additional challenges for spindle identification due to its unique signal characteristics and recording environment.

This study introduces a novel, precise, and automatic spindle detection method for iEEG using a dual-head architecture to enhance performance, robustness, and ease of use. Our approach achieves a detection F1 score of 0.67 on a challenging iEEG dataset and 0.69 on the publicly available scalp EEG DREAMS dataset. Compared to existing methods such as SUMO, A7, and YASA, our model demonstrates superior performance in detecting, segmenting, and characterizing sleep spindles. This model contributes to open science and advances automated sleep spindle classification in iEEG. This will advance the development of more precise diagnostic and research tools and facilitate a deeper understanding of the role of sleep spindles in cognitive processes and neurological health.

## 1 Introduction

Sleep spindles (Figure 1) are brief bursts of oscillatory brain activity characterized by their distinct waxing and waning shape, which usually occur in the sigma frequency range of 11–16 Hz [13]. They are a prominent feature observed in sleep electroencephalographic (EEG) recordings, mainly during non-rapid eye movement (NREM) stage N2 [13]. Spindles are thought to reflect a state of thalamocortical synchrony, and appear to play a crucial role in various brain functions [13, 9].

**Figure 1:**
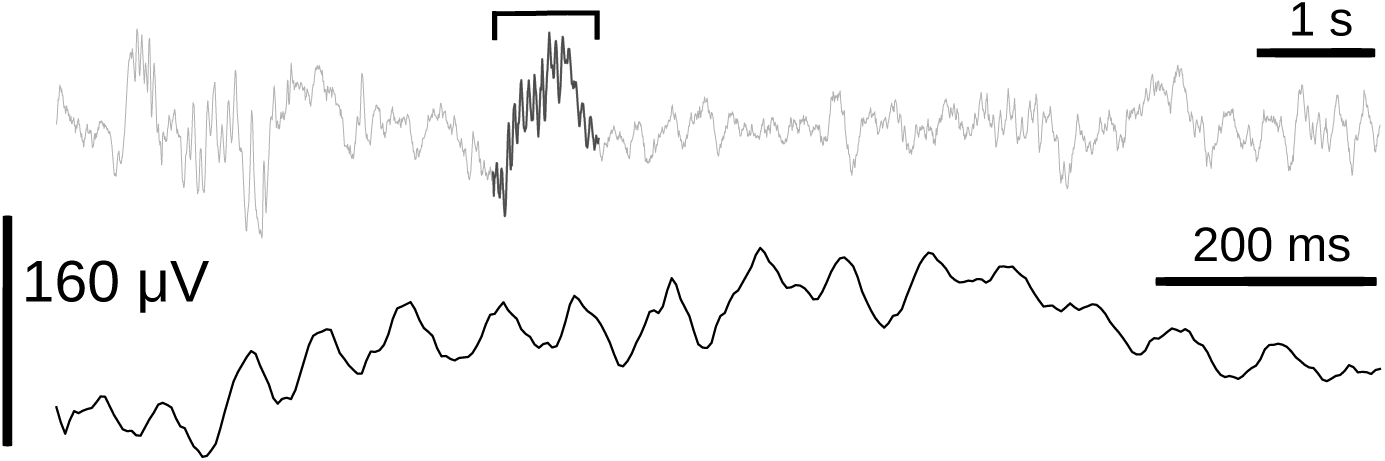
Sleep spindle example. Intracranial local field potential (LFP) activity recorded at 250 Hz from the left anterior nucleus of thalamus (ANT) during NREM II (N2) sleep in a patient from our epilepsy monitoring unit. A 10-second segment is shown, with a 1.2-second sleep spindle highlighted. The spindle is enlarged below to illustrate its oscillatory nature. With a duration of approximately *D ≈* 1.2, s and 15 observable peaks, its frequency is estimated to be around 12.5Hz.

Their consistent presence and characteristics within an individual from night to night are an essential feature of sleep architecture. In addition, spindles are believed to contribute significantly to processes such as synaptic plasticity and memory consolidation, making them integral to learning and optimal cognitive functioning. [17] Variations in spindle density, amplitude, and duration occur during brain development and can provide insights into neurological functioning and integrity of brain circuits involved in learning and memory. Clinically, spindle activity abnormalities are associated with various neurological and psychiatric disorders, highlighting their potential as biomarkers for epilepsy, schizophrenia, autism, and neurodegenerative diseases. [6, 25].

In standard clinical settings in sleep medicine, experts visually detect spindles as a specific pattern of sleep microarchitecture to identify non-REM stage N2 sleep. Spindles are not labeled or used for any other quantification of sleep evaluation in clinical settings. Labeling each spindle during sleep is enormously time-consuming and challenging, and is not scalable in most clinical or research settings. On the other hand, automatic precise detection and quantification of sleep spindles is crucial in research and could further pave the way to clinical practice application if automated tools are shown to be reliable and become readily available. Subtle variations in spindle frequency and patterns can provide important insights into brain health and function, and might be useful toward some future clinical applications, such as predicting early-stage sleep or neuropsychiatric disease development and assessing longitudinal disease course and treatment outcomes. Accurate and automated spindle detection would enable detailed analysis and quantification of spindle characteristics, such as their frequency, amplitude, and distribution in different regions of the brain. Furthermore, precise detection is essential for studying the role of spindles in cognitive processes, as even small changes in spindle dynamics can reveal changes in brain networks underlying learning, memory, and overall cognitive performance [12, 15, 5].

Most sleep spindle analyses have been performed using scalp EEG sleep recordings. However, recently, with the development of technology and advances in neurological disorder diagnostics, intracranial EEG (iEEG) offers an unique opportunity to study brain network dynamics, including sleep spindle analysis. iEEG includes deeper brain structures and offers higher spatial sampling of distinct topographic neuroanatomical regions underlying scalp EEG detected oscillations and neural circuit functioning. [22, 32, 27, 39]. Direct analysis of specific brain regions by iEEG allows for more detailed and localized monitoring of neuronal networks essential for identifying subtle activity patterns, such as the intricate dynamics of sleep spindles and their interaction between cortical and deep brain structures. [31, 21] Regrettably, there are relatively few tools designed for the automated detection, parameterization, and quantification of sleep spindles. The majority of existing tools have been developed and evaluated exclusively using public datasets comprised of scalp EEG recordings, resulting in limited generalizability and applicability to iEEG data.

Our study focuses on advancing spindle detection using iEEG local field potentials to better understand the role of spindles in brain function and health. To overcome the aforementioned insufficiencies, we developed a generalized, open, deep learning method that excels in accuracy and efficiency. We demonstrate its superior performance metrics compared to existing state-of-the-art models for both scalp and iEEG data, where our approach achieves the highest F1 scores across datasets, highlighting its robustness and practical utility. By automating and refining spindle detection methods, we aim to improve future research and clinical outcomes and provide a deeper understanding of the role of sleep spindles in brain function and health. We publish our methods in a GitHub repository (https://github.com/CaptainTrojan/mayo spindles), including Python codes and trained models, to facilitate open science and brain research. Access to these resources, along with an MIT License, will allow researchers to reproduce our analyses and explore new applications of our models.

## 2 Related Work

Various methods have been used to detect sleep spindles, each having a unique approach, strengths, and weaknesses. To date, all methods have been developed for scalp EEG data to our knowledge. Currently, no previous methods have been published to address sleep spindle detection from iEEG recordings.

The A7 algorithm [24] is a feature-based machine learning (ML) algorithm designed to emulate the expert human scoring of sleep spindles using a single EEG channel, specifically C3-A2. It works by analyzing four key parameters: absolute sigma power, relative sigma power, sigma covariance, and sigma correlation, all of which are computed within a sliding window of 0.3 seconds. Spindle detection is based on identification of candidate spindles exceeding predefined thresholds for these parameters. The A7 algorithm has shown a precision of 74%, recall of 68%, and an F1 score of 0.70 when evaluated against a gold standard data set of human expert scored spindle labels. Its balance between sensitivity and precision makes it a reliable tool for automated spindle detection. [24]

YASA [35], an open-source feature-based ML tool for automated sleep staging, provides a sophisticated method for spindle detection by leveraging a variety of metrics. It calculates relative power, moving correlation, and root mean square (RMS) from band-filtered EEG data. YASA’s approach includes thresholding these metrics to identify potential spindle events and applying duration and proximity filters to refine detections. The algorithm also aggregates spindle detections across multiple channels to improve reliability. YASA is notable for its speed in processing EEG signals, although its precision and recall performance can substantially vary between different datasets.

SUMO [16] utilizes a deep neural network inspired by the U-Net architecture to detect sleep spindles. The model processes preprocessed EEG segments to generate probability maps that indicate spindle likelihood. These maps are then thresholded to identify the spindles. SUMO achieved F1 scores of 0.75 in younger individuals and 0.65 in older individuals. The training of the SUMO model involves early stopping and the Adam optimizer to enhance accuracy, making it a robust approach to spindle detection.

Wendt et al. [38] developed an ML algorithm that integrates bandpass filtering with envelope detection and decision fusion from two detectors placed at the central electrodes (C3-A2) and occipital electrodes (O1-A2). Their method filters the EEG signals to isolate the spindle frequency range and uses the resulting envelopes to detect spindles by analyzing their local extrema. A decision fusion approach combines the outputs from both detectors, enhancing the accuracy and robustness of spindle detection by addressing various factors affecting detection performance.

McSleep [30] focuses on multichannel spindle detection through a sparse low-rank optimization approach. This method assumes that the EEG signal comprises transient and oscillatory components, with the oscillatory component modeled as low-rank arrays. By separating these components using a convex optimization problem, McSleep detects spindles across multiple channels in a single run, demonstrating efficiency and minimal parameter tuning.

MUSSDET [23] employs a support vector machine (SVM) to detect sleep spindles in segmented EEG epochs. The method includes feature selection through correlation-based filtering to reduce dimensionality and optimize SVM parameters for classification. The detector is designed to be consistent and robust across different datasets, avoiding the need for extensive parameter adjustments and achieving reliable spindle detection.

Martin et al. [26] describes a method for automatic detection of sleep spindles in artifact-free NREM sleep epochs from scalp EEG recordings. The algorithm processes data from multiple para-sagittal scalp derivations (left: Fp1, F3, C3, P3, O1; right: Fp2, F4, C4, P4, O2) by first applying a bandpass filter between 11 and 15 Hz to isolate spindle frequencies. A linear-phase Finite Impulse Response (FIR) filter is used to ensure zero-phase distortion, followed by the calculation of the root mean square (RMS) of the filtered signal with a 0.25-second time window. The resulting RMS values are thresholded at the 95th percentile to identify spindle events. A spindle is confirmed when at least two consecutive RMS points exceed this threshold, with a minimum duration criterion of 0.5 seconds.

Wamsley et al. [36] uses a wavelet-based method to detect discrete sleep spindles by applying an 8-parameter complex Morlet wavelet to the raw EEG signal. The method performs a time-frequency transformation to extract spindle-related features in the 10 Hz to 16 Hz frequency range. A rectified moving average is computed using a 100-millisecond sliding window to threshold the wavelet signal. A spindle is identified when the wavelet signal exceeds 4.5 times the mean amplitude of all artifact-free epochs for a duration of at least 400 milliseconds.

Bódizs et al. [2] detects sleep spindles by first calculating the amplitude spectrum of NREM sleep EEG signals and identifying the frequency boundaries for slow and fast spindles. The signal is then bandpass-filtered, rectified, and smoothed to extract an envelope, which is compared to amplitude criteria to detect spindle events lasting more than 0.5 seconds.

Gais et al. [14] detected sleep spindles by filtering the EEG within the 12–15 Hz range, calculating the root mean square (RMS) of 100 ms intervals, and identifying when the RMS power exceeds a threshold of 10 µV for 0.5 to 3 seconds. This algorithm accurately identified approximately 95% of spindles compared to visual scoring on internal data used in their study. Devuyst et al. [11] detects pseudo K-complexes by extracting various features from the EEG signal and applying a multi-level thresholding approach to assess the likelihood of an EEG event being a K-complex, instead of simply rejecting or accepting an event based on fixed thresholds. The algorithm computes a likelihood score for each probable K-complex event using fuzzy thresholds defined by a pre-established “thresholding curve” for each feature (e.g., duration, voltage, waveform morphology, and others). This method, while originally designed for K-complex detection, can be adapted for spindle detection by modifying the feature set and thresholding curves specific to spindles, allowing for a flexible and accurate detection system.

The DETOKS algorithm [29] detects sleep spindles and K-complexes by separating the EEG signal into two components: a low-frequency transient component and an oscillatory component. The oscillatory component, which is used for spindle detection, is bandpass filtered (11.5-15.5 Hz) and then processed using the Teager-Kaiser Energy Operator (TKEO) to define a binary signal. Spindles are detected if their duration is between 0.5 and 3 seconds. Similarly, the TKEO is applied to the low-frequency component for K-complex detection, with a thresholding method to define binary events. Both spindle and K-complex detections are subject to duration constraints of at least 0.5 seconds. This approach outperforms existing methods, showing higher F1 scores for both sleep spindle and K-complex detection.

Chen et al. [7] introduced a Deep Neural Network (DNN) framework for spindle detection that combines deep features with entropy measures of EEG epochs. Their approach uses a compact Convolutional Neural Network (CNN) with spatial pyramid pooling and focal loss to adapt to varying spindle durations and improve detection accuracy. The DNN framework outperforms traditional methods by enhancing the representation of complex spindle waveforms, leading to more accurate detection of spindle onsets and offsets.

In conclusion, numerous techniques have been employed to detect sleep spindles, each employing distinct strategies and technologies. These techniques can generally be divided into two primary branches: feature engineering with machine learning, utilizing bandpass filtering and thresholding, and deep learning approaches.

### Feature Extraction and Thresholding Approaches

Methods such as the A7 algorithm [24] utilize specific parameters such as absolute sigma power, relative sigma power, sigma covariance, and sigma correlation, operating on a single EEG channel. Although effective, these methods are heavily based on predefined thresholds and parameters. Similarly, YASA [35] uses metrics such as relative power, moving correlation, and root mean square (RMS), focusing on thresholding and duration filtering, and aggregates spindle detections across multiple channels. Wendt et al. [38] combine bandpass filtering with envelope detection and decision fusion from two detectors to enhance robustness. McSleep [30] employs sparse low-rank optimization for multichannel detection, aiming at efficiency with minimal parameter tuning.

### Deep Learning Approaches

In contrast, modern deep learning ML methods like SUMO [16] leverage neural network architectures to detect spindles, utilizing advanced models inspired by the U-Net architecture. SUMO processes preprocessed EEG segments to generate probability maps for spindle detection, achieving high F1 scores. Chen et al. [7] introduced a Deep Neural Network (DNN) framework that integrates deep features with entropy measures, using a compact Convolutional Neural Network (CNN) with spatial pyramid pooling and focal loss to adapt to spindle variations, thereby improving detection accuracy.

### Our Contributions

While existing methods have made significant strides, they often rely on older architectures or approaches that may not fully leverage recent technological advancements. Our method differentiates itself by utilizing modern deep-learning architectures and integrating two distinct heads to enhance performance and representation. This innovative approach builds on the strengths of contemporary neural network models and introduces improvements such as multitask inference, internal ensembling, and advanced feature extraction techniques. The dual-head architecture allows for a more nuanced representation of spindle characteristics and better overall performance, as a single shared feature extraction backbone feeds into two separate output “heads,” each performing a different prediction task (detection and segmentation). By combining these advanced strategies, our method addresses some of the limitations of existing approaches and offers a more robust solution for both research and clinical applications. Our method is unique in generalizing sleep spindle detection to EEG and iEEG data.

## 3 Method

### 3.1 Datasets

We evaluated our spindle detection algorithm on two datasets: the publicly available DREAMS database [10] and the proprietary Bioelectronics Neurophysiology and Engineering Lab (BNEL) intracranial EEG (iEEG) dataset. The BNEL iEEG dataset was collected at the Mayo Clinic and is available from the authors upon reasonable request. Additional details about the BNEL lab can be found at https://www.mayo.edu/research/labs/bioelectronics-neurophysiology-engineering/overview.

#### 3.1.1 DREAMS Dataset

The DREAMS dataset [10] is a comprehensive collection of polysomnographic recordings compiled as part of the DREAMS project, funded by Région Wallonne, Belgium. The DREAMS databases include a variety of annotated sleep data collected from healthy subjects and patients with various pathologies.

The DREAMS Sleep Spindles Database contains eight 30-minute segments from central EEG channels, derived from full-night PSG recordings of subjects aged 41 to 53 years, except for a single excerpt involving a 31-year-old male participant [10]. These excerpts are annotated by two independent experts for sleep spindles. Notably, expert 1’s spindle counts were capped after 1000 seconds, which may affect direct comparison of spindle counts.

The recordings were obtained using a 32-channel polygraph (Brainnet TM System by MEDATEC, Brussels, Belgium) and stored in the European Data Format (EDF). The data set is publicly available to facilitate future research and performance comparisons. [10]

Considering that the dataset contains annotations from two annotators and recognizing that an annotator is significantly more likely to overlook spindles rather than misclassify baseline EEG as a spindle, we defined the ground truth as the union of the labels provided by both annotators. This approach follows the preferred method established in the literature [7], [30], as the DREAMS dataset does not specify an alternative for synthesizing ground truth from multiple annotators.

#### 3.1.2 BNEL Mayo Clinic EMU Data (BNEL iEEG)

This dataset comprises simultaneously scalp EEG and iEEG data from a chronically implanted brain stimulator. Data were recorded in the Epilepsy Monitoring Unit (EMU) setting in a cohort of patients with drug-resistant temporal lobe epilepsy. Data were provided by the Bioelectronics Neurophysiology and Engineering lab of Mayo Clinic in Rochester, Minnesota. The dataset includes iEEG data from four epileptic patients [34]; specifically:

- Two nights of data from Patient 1 (female, 57 years old)
- One night of data from Patient 2 (female, 20 years old)
- Two nights of data from Patient 3 (female, 35 years old)
- One night of data from Patient 4 (female, 41 years old)

Each patient has multiple electrodes implanted in bilateral anterior nucleus of the thalamus (ANT) and bilateral hippocampus (HPC). The implanted device sprovides a continuous stream of four channels of iEEG data: *ANT_L_* (left thalamus), *ANT_R_* (right thalamus), *HPC_L_* (left hippocampus) and *HPC_R_*(right hippocampus). The recordings were annotated by an expert (V.V.). Each channel was individually annotated for scalp EEG and iEEG spindles. During post-processing, the channels were isolated to treat each channel separately as a single-channel 1D detection task. Each annotated spindle retains its associated channel information, allowing us to evaluate the prediction quality for each channel independently.

### Experimental Protocol

This research involving human participants was conducted with approval from a Food and Drug Administration investigational device exemption (IDE-G180224) and the Mayo Clinic institutional review board (IRB: 18-005483 “Human Safety and Feasibility Study of Neurophysiologically Based Brain State Tracking and Modulation in Focal Epilepsy”). It is registered at https://clinicaltrials.gov/ct2/show/NCT03946618. Participants provided informed consent according to IRB and FDA guidelines. Four individuals with drug-resistant mesial temporal lobe epilepsy (mTLE) consented to be implanted with the investigational Medtronic Summit *RC* + *S^T^ ^M^* device (INSS) [34, 20], featuring four contact leads, targeting bilateral HPC and ANT. For more information related to surgical and clinical aspects, refer to our previously published studies [34, 20]. We collected concurrent polysomnography, EEG, and iEEG data in a hospital environment. The polysomnogram was scored by a board certified sleep specialist (EKS). The overnight hospital and long-term study were part of a broader clinical trial testing brain stimulation for treatment of drug resistant temporal lobe epilepsy [34, 20].

### Intracranial EEG Data Acquisition

Data from intracranial EEG was continuously gathered via wireless streaming to a tablet computer at a sampling rate of either 250 Hz or 500 Hz, following previous methodologies [20, 34, 28]. Data originally collected at 500 Hz was filtered and downsampled to 250 Hz using a 101st order anti-aliasing finite response zero phase shift filter with a cutoff at 100 Hz. The impedance between the implanted device and electrode contacts remained below 2 *k*Ω for all contacts throughout the experiments. We analyzed iEEG data from three consecutive days of hippocampal (HPC) iEEG recording without ANT DBS to differentiate the Awake, rapid-eye-movement (REM), and non-REM (N2 & N3) sleep stages. Sleep stages were scored by experts for all patients, as depicted in Figure 2.

**Figure 2:**
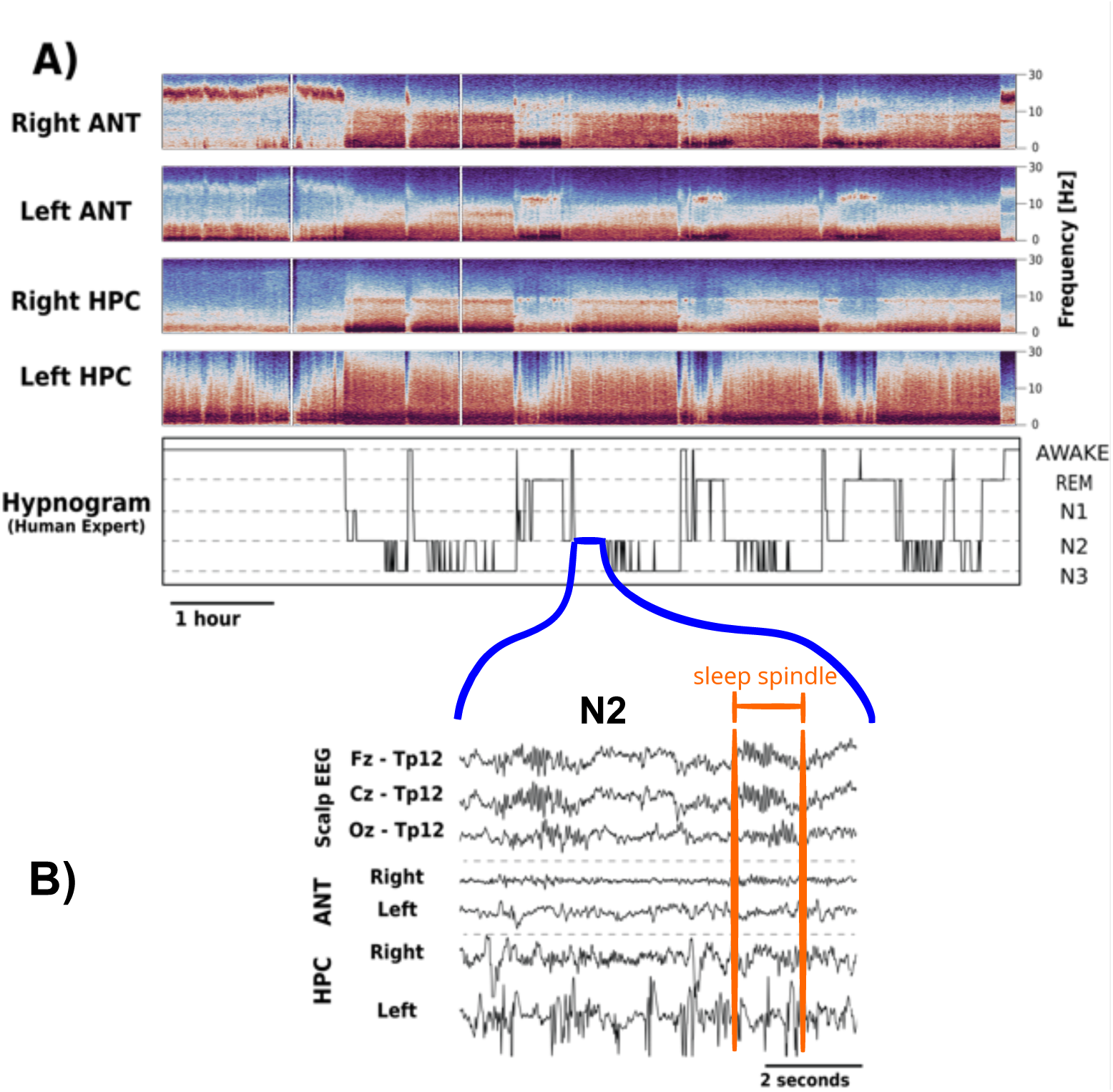
Variations in intracranial EEG (iEEG) signals can be observed across various behavioral states, such as Awake, REM, N2, and N3. A) The iEEG spectrograms, obtained from the left and right anterior nuclei of the thalamus (ANT) and hippocampus (HPC), illustrate changes linked to Awake, REM, and non-REM states. Notable differences exist in the iEEG signals from the left and right ANT and HPC, particularly concerning epilepsy’s electrophysiological markers, for instance, the left HPC exhibits a higher frequency of interictal epileptiform discharges. B) Concurrent recordings of scalp-EEG (positions Fz, Cz, and Oz referenced to A2) and iEEG (bipolar electrodes on Left ANT, Right ANT, Left HPC, Right HPC) for the N2 stage, where sleep spindles are annotated by a specialist.

### Sleep Scoring Using Scalp EEG

For the expert visual scoring of sleep stages, all PSG records were filtered through a band-pass of 0.3 to 75 Hz, incorporating a 60 Hz notch filter using sixth-order zero-phase-shift Butterworth filters. Electrodes were located according to the standard 10-20 system, including eye (electrooculogram, EOG) and chin (electromyogram, EMG) electrodes for analyzing eye movements and muscle activity crucial for accurate REM sleep state determination. The visual scoring of sleep stages was manually conducted by an expert reviewer (EKS) following the American Academy of Sleep Medicine’s scoring guidelines [33], utilizing the CyberPSG software by Certicon a.s. Wakefulness was identified by eye blinks in the frontal scalp and eye leads, combined with a posterior dominant alpha rhythm (8–12 Hz) covering more than 50% of the epoch. Stage N2 sleep was marked by low-frequency delta and theta frequency EEG waves predominant in 30 second epochs, along with the presence of K complexes or sleep spindles. N3 sleep, or slow-wave sleep, was noted when at least 20% (6 seconds) of 30-second epochs showed high-voltage (*>* 75*µV*), delta (0.5 – 4 Hz) activity in frontal EEG leads. REM sleep was classified based the emergence of rapid eye movements (¡500 msec) on electrooculogram (EOG) derivations associated with reduction in submentalis electromyography (EMG) tone and low-amplitude mixed-frequency EEG activity. This methodology aligns with prior studies and typically utilized clinical and research sleep stage scoring standards [18, 19, 3]. In entirety, an expert reviewer annotated a total of 12,182 sleep epochs using the PSG recordings. The mean distribution of sleep stages was as follows: Awake - 29.4%, N1 - 4.8%, N2 - 37.7%, N3 - 15.7%, and REM - 12.32%. Figure 3 illustrates the block diagrams for the experimental setup and data pipelines.

**Figure 3:**
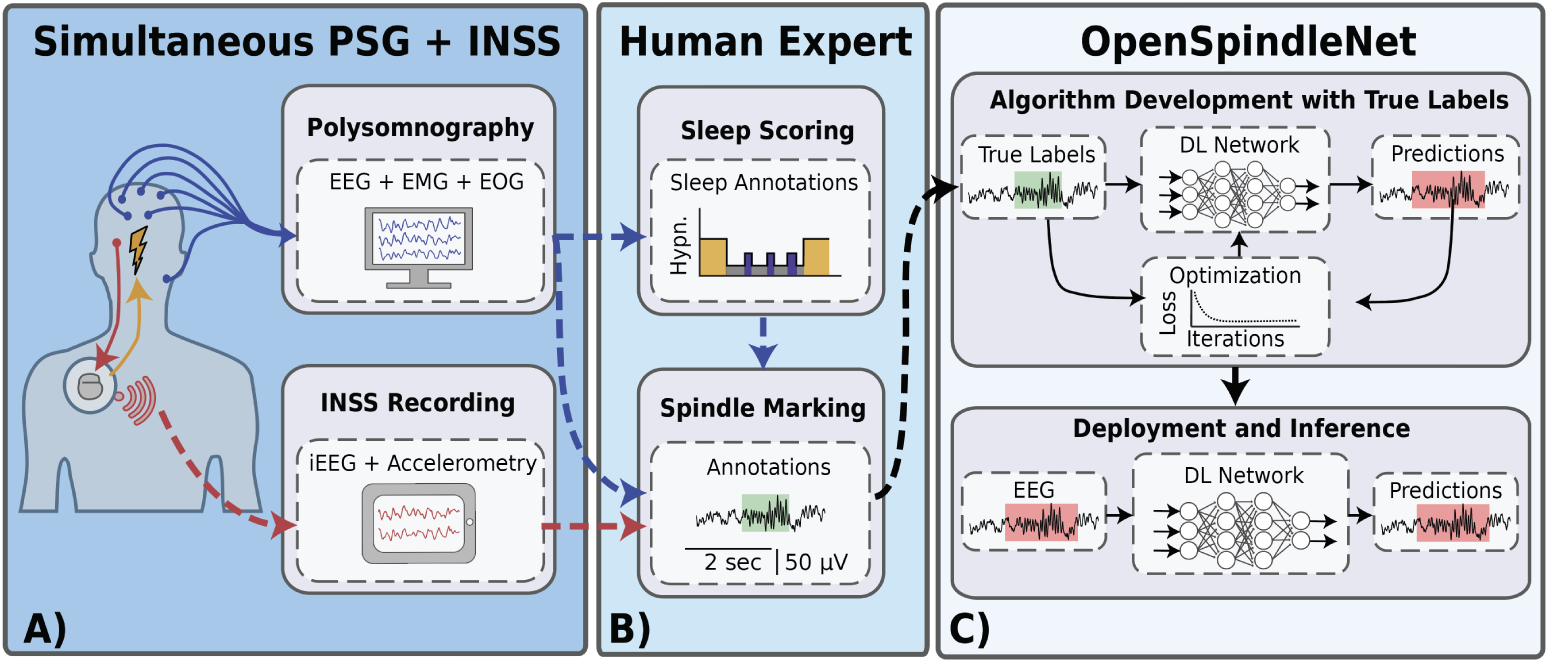
Block-diagram of the experimental setup and data pipelines of sleep data and spindle data acquisition, labeling, and classification. A) Schematic representation of the concurrent scalp electroencephalography (EEG) and intracranial EEG (iEEG) acquisition setup. B) Workflow illustrating recording of iEEG using implantable neural stimulation sensing (INSS) device and scalp EEG with polysomnography (PSG). The PSG data scored by expert clinicians, served as a gold standard for sleep stage classification, subsequently applied to both scalp and iEEG recordings. Independent expert annotation of sleep spindles was performed on both scalp and iEEG data, providing labeled datasets for model training and evaluation. C) Diagram depicting the data partitioning strategy for model development and evaluation. The dataset was divided into training, validation, and testing sets, ensuring sequential model training to prevent data leakage. The final stage involved deploying the trained OpenSpindleNet model to unseen data. Model performance was assessed by direct comparison with expert-annotated sleep spindle labels.

### Data Processing

The iEEG and PSG recordings were stored in the Multi-Scale Electrophysiology Format version 3 (MEF) [4] and imported into Python using a Python library “pymef” available on GitHub (https://github.com/msel-source/pymef). The gold standard annotations for sleep scoring were exported from the CyberPSG software and imported into Python using the software package Behavioral State Analysis toolbox (BEST, https://best-toolbox.readthedocs.io/en/latest/index.html#).

### 3.2 Experimental Setup

To develop and evaluate our spindle detection model, we followed a standard machine learning workflow. Each dataset was split into training, validation, and test sets in ratio 70/15/15. The resulting split sizes are shown in Table 1. The model was trained on the training set, and hyperparameters were selected in order to optimize performance on the validation set. The training procedure is described in Section 3.4 and the configuration that we identified as optimal shown in Section 3.4.5.

**Table 1:** Sizes of BNEL iEEG and DREAMS. The table presents the number of segments and ground truth spindles for each dataset split (train, val, test) for both BNEL iEEG and DREAMS datasets.

|  | BNEL iEEG |  |  | DREAMS |  |  |
| --- | --- | --- | --- | --- | --- | --- |
|  | Train | Val | Test | Train | Val | Test |
| N<sup>o</sup> segments (30s) | 425 | 91 | 92 | 167 | 35 | 37 |
| N<sup>o</sup> spindle labels | 682 | 159 | 189 | 415 | 72 | 91 |

After selecting the best-performing configuration, the model was evaluated on the test set to assess final performance in an unbiased manner, ensuring the evaluation data is independent of all data selected for training and optimization, while remaining identically distributed to data expected to be encountered in deployment. The final results measured on the test set of both intracranial (BNEL iEEG) and scalp data (DREAMS) are summarized in Section 4.

We opted for a fixed train/validation/test split rather than cross-fold validation because, for our internal dataset, the validation and test sets were meticulously cleaned and curated, while the training set remained relatively noisy. This strategy enabled us to properly focus our labeling efforts and assess the model’s performance on clean, high-quality data, offering a more accurate measure of its generalization. Cleansing the test data also ensured that the evaluation of other state-of-the-art methods was fair and unbiased.

### 3.3 Spindle detector

We have chosen to employ a deep-learning approach for our spindle detection model. Several advantages of deep learning methods that drove this choice include that they are inherently noise-resistant, robust under various conditions, and capable of directly processing raw input signals. This eliminates the need for extensive domain knowledge typically required for manual feature extraction. Additionally, the use of deep learning makes the model task-agnostic, meaning that the same or similar architectures can be effectively applied to different detection tasks, such as spindle, inter-ictal epileptiform activity, or K-complex detection.

#### 3.3.1 Model inputs

The recordings are divided into 30-second blocks. Each block is then processed independently by resampling the signal to 250 Hz, which results in a vector with 7500 voltage values.

The signal from each block is processed in two distinct and parallel pathways. First, the signal is standardized to have a mean of 0 and a standard deviation of 1. Critically, in the second pathway, a continuous wavelet transform (CWT) is applied directly to the original, non-normalized signal. This intentional separation ensures that the CWT retains the raw amplitude information of the unprocessed signal, ensuring the baseline is not deviated and the amplitudes are unaltered. The CWT generates a scalogram with 15 frequency bands spanning approximately 9 to 18 Hz, a range chosen to capture spindle activity with a small additional margin. The scalogram is represented as a matrix of shape (15, 7500), where each row corresponds to a specific frequency band.

The standardized signal (normalized to zero mean and unit variance) and the scalogram (derived from the raw, unaltered signal) are then concatenated along the first axis, resulting in a combined input with a shape of (16, 7500). This dual-input approach leverages both normalized temporal features and the unprocessed spectral characteristics of the signal. An example of these inputs is shown in Figures 4 and 5.

**Figure 4:**
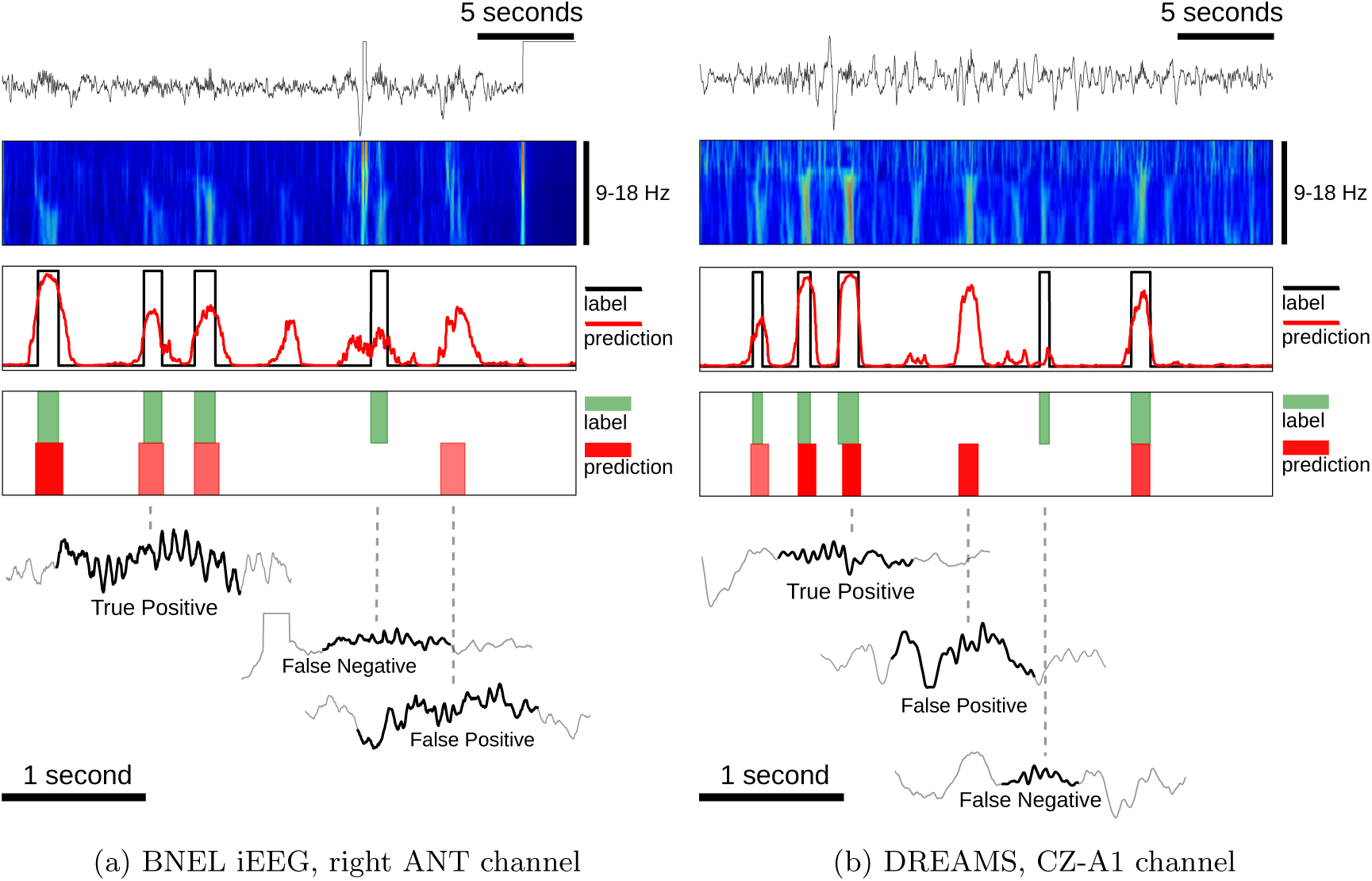
Model input and output predictions. The top panels display the normalized input signal and its corresponding scalogram. The bottom panels show the outputs from the model’s segmentation and detection heads. In the segmentation plot (top), the black line denotes the ground truth, while the red line indicates the model’s predicted segmentation confidence (ranging from 0 to 1). In the detection plot (bottom), green blocks mark ground truth spindle annotations, and red blocks represent the model’s predictions with confidence *>* 0.5 and intersection-over-union (IoU) *<* 0.3. Each plot highlights examples of a true positive (correct spindle detection), false positive (incorrect spindle detection on baseline signal), and false negative (missed spindle).

**Figure 5:**
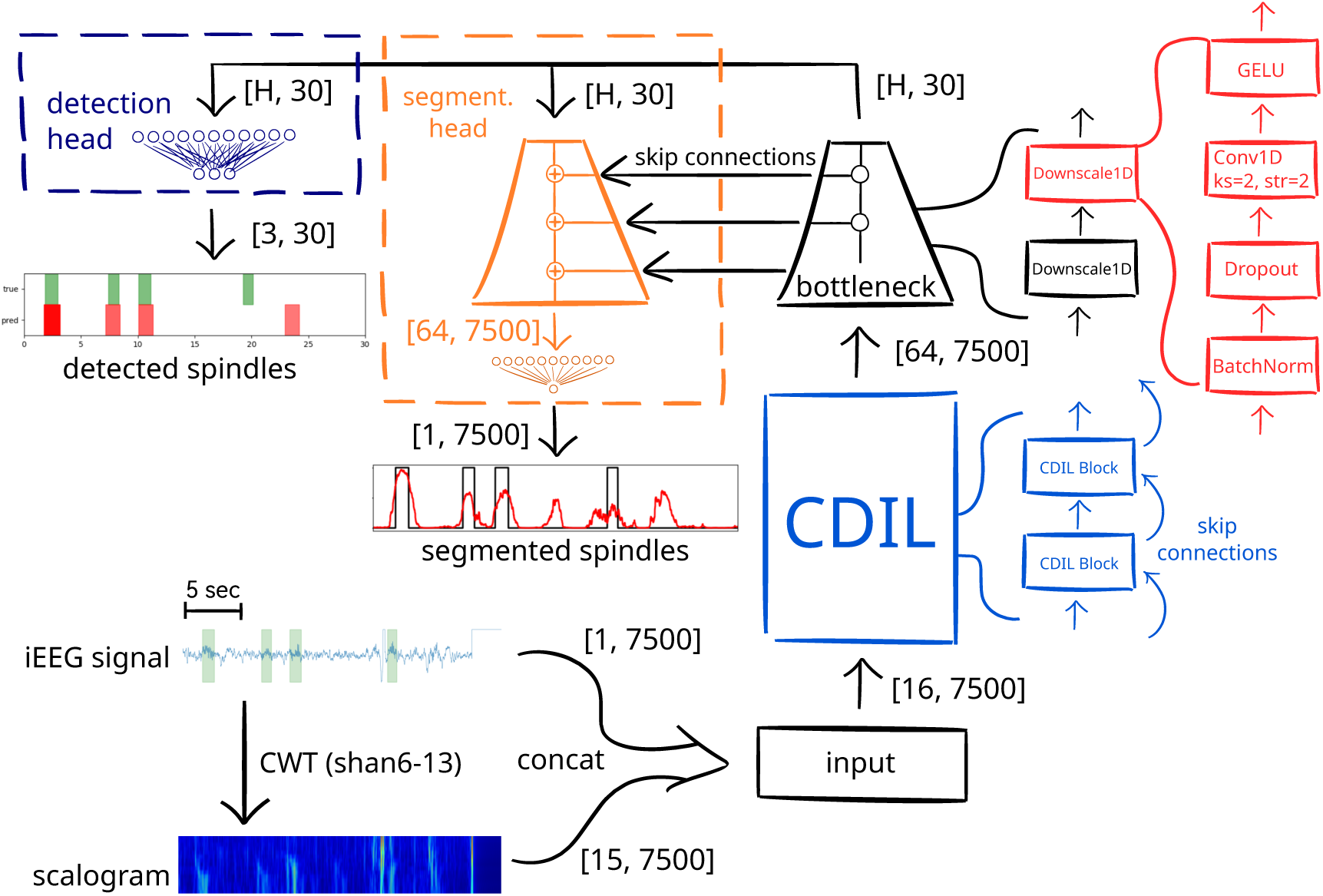
The model architecture. A shared bottleneck variant, see Section 3.3.3 and Figure 6 for details. The flow of tensors between different components of the network is annotated with the corresponding tensor shapes. For example, the Circular Dilated Convolution (CDIL) output, which serves as the bottleneck input, has shape [64, 7500].

#### 3.3.2 Architecture

The proposed model is a specialized 1D Convolutional Neural Network (CNN) designed with two distinct prediction heads: one for spindle detection and another for spindle segmentation. The overview of the architecture is visualized by Figure 5.

### Backbone

The backbone of the model is based on Circular Dilated Convolution (CDIL) [8], an efficient 1D Convolutional Neural Network (CNN) architecture that leverages circular padding, dilated convolution kernels, and residual connections. This design allows CDIL to capture context over longer sequences more effectively than traditional methods, where the context grows exponentially with layer depth rather than linearly. The backbone processes input matrices of shape (16, 7500), producing contextual embeddings of shape (64, 7500), where each time-step in the input is represented in a richer, high-dimensional space.

### Bottleneck

To train the model for both detection and segmentation, we utilized a bottleneck block that down-scales (compresses) the backbone’s output. This compression reduces the sequence from 7500 embeddings to 30 embeddings using a chain of 1D CNN layers (with batch normalization and dropout, see Figure 5). Now, instead of having an embedding for each time-step of the signal, we have one for each second of the signal. Since spindles typically last around one second, this approach helps the network to focus on the presence of spindles within each second-long interval. The output of the bottleneck block is a matrix of shape (H, 30), where H represents the “hidden size”: a hyperparameter optimized during the training process. Then, each prediction head separately processes this compressed representation. The bottleneck also retains intermediate representations after each down-scaling layer, which are used for residual connections during the up-scaling process in the segmentation head (see below).

### YOLO-like Detection Head

The detection head processes the bottleneck output to produce a tensor of shape (3, 30) through a small multi-layer perceptron (MLP) that operates along the channel dimension (H *→* 3). The features are then passed through a sigmoid activation function to constrain the values between 0 and 1. Hence, we can think of the result as the model predicting three features (ranging from 0 to 1) for each second of the input signal:

1. spindle existence confidence (indicating whether a spindle is present),
2. spindle center offset relative to the interval’s center (ranging from 0 = spindle center is at the interval start, to 1 = spindle center is at the interval end), and
3. spindle length (ranging from 0 to 1, linearly interpolated to 0.5 *−* 2 seconds, which is the typical spindle duration).

These predictions are then converted to start and end points and rounded to the nearest integer indices. Finally, to mitigate overlapping detections during inference, we apply the Non-Maximum Suppression (NMS) filter with an intersection-over-union threshold of 0.3. See Figure 4 for an example prediction.

### Segmentation Head

The segmentation head up-scales the bottleneck output using the same structure as the bottleneck but with transposed convolutions (also known as reverse convolutions or de-convolutions). The input is up-scaled from (H, 30) back to (H, 7500), then reduced to (1, 7500) through another MLP along the channel dimension (H *→* 1). Furthermore, each up-scaling layer adds the corresponding intermediate feature from the bottleneck, combining both low and high-level features from the CNN layers for the final MLP. The resulting tensor is passed through a sigmoid layer to produce confidence scores for each time-step, indicating the likelihood that a given time-step is part of a spindle. As a post-processing step, we apply a median filter to the resulting segmentation with window size 11 to smooth out the segmentation map. See Figure 4 for an example prediction.

#### 3.3.3 Head Variants

We developed three different versions of the model head to assess the value of using a dual-head configuration. The choice of head variant essentially acts as a hyperparameter that must be tuned. The optimal variant will be selected based on its performance on the validation split during training. A diagram of each head variant can be seen in Figure 6.

**Figure 6:**
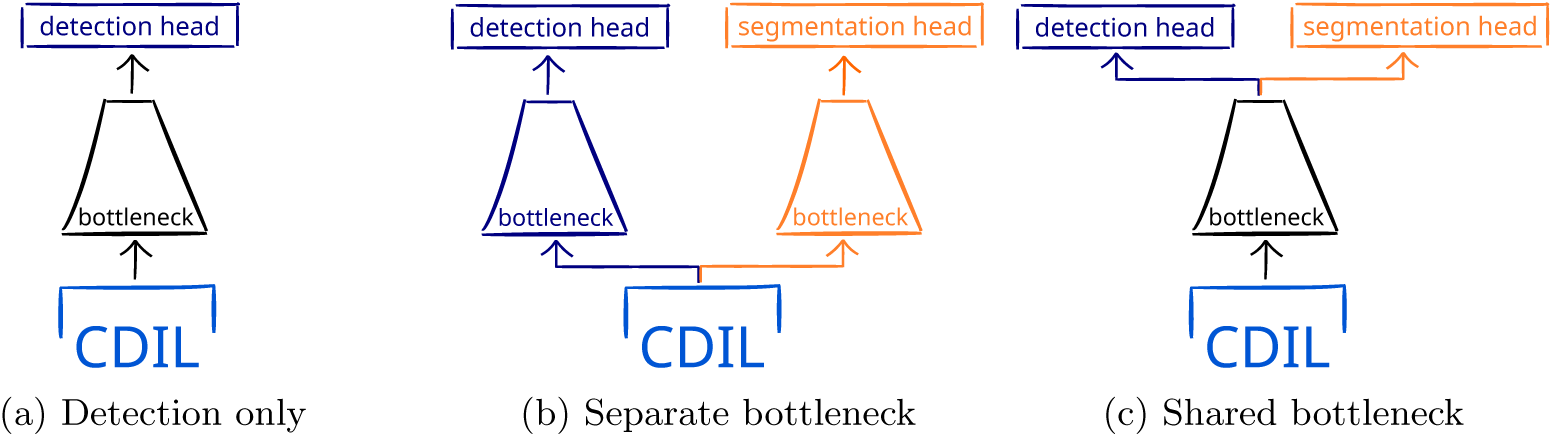
Three different head variants were considered during the architecture design.

##### Detection Only

This variant is designed solely for the detection task, excluding the segmentation head altogether.

##### Separate Bottleneck

In this variant, both the detection and segmentation heads are included, with the bottleneck duplicated for each head. Essentially, after the backbone, there are two separate bottlenecks trained independently. This variant has the highest number of trainable parameters.

##### Shared Bottleneck

This variant leverages a single bottleneck shared between the detection and segmentation heads, which means that they both use the same set of weights. As a result, it has fewer parameters than the separate bottleneck variant while still including both heads.

### 3.4 Training

This section offers a comprehensive overview of the training methodology used to develop our spindle detection model.

#### 3.4.1 Loss calculation and evaluation

The loss function for training the model integrates three components to address both spindle detection and segmentation. First, the loss in detection probability, *L*_det prob_, is calculated using binary cross-entropy (BCE) between the predicted and actual values to determine the presence of the spindle (center) in each of the 30 intervals (1s long). Second, for intervals where spindles are present, the detection parameters loss, *L*_det params_, measures the mean squared error (MSE) between the predicted and true spindle center offsets and durations. Finally, the segmentation loss, *L*_seg_, uses BCE to compare the predicted segmentation map with the ground truth. The “detection only” head variant omits the last component.

The overall loss is the sum of these three components:

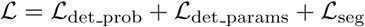

The following are the metrics we used to evaluate the detection and segmentation performance.

##### Detection F1 Score (@ IoU=0.3)

The detection F1 score quantifies the balance between precision and recall in spindle detection by calculating their harmonic mean, providing a single metric that captures the trade-off between false positives (FP) and false negatives (FN). Specifically, the detection F1@IoU=0.3 evaluates this score by treating predictions as true positives (TP) if their Intersection over Union (IoU) with ground-truth labels is 0.3 (30% overlap) or higher. This metric effectively measures the model’s ability to achieve both precision and recall when predictions overlap with ground-truth labels by a significant amount [7]. We set the confidence threshold at 0.5 to classify predictions as either positive or negative.

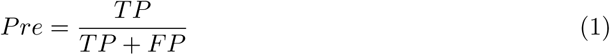

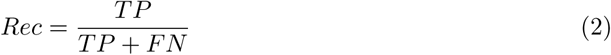

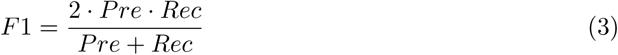

##### Segmentation F1 Score

To evaluate sleep spindle detection performance, we employed both a detection-based F1 score and a segmentation-based F1 score. The segmentation F1 score, also referred to as the per-timestep or by-sample F1 score [30], provides a more granular assessment by considering individual samples (at 250 Hz) within each 30-second epoch. Unlike the detection-based F1 score, which evaluates entire spindle events, the segmentation F1 score compares the model’s classification of each sample (spindle or non-spindle) to the corresponding ground truth label. This per-sample evaluation allows for a more detailed analysis of performance, particularly regarding the temporal accuracy of spindle boundaries. True positives, false positives, and false negatives are calculated based on these per-sample classifications. Figure 7 visually illustrates the distinction between these two metrics. Consistent with prior work [38, 30, 23, 29], and given its sensitivity to boundary accuracy, we selected the segmentation F1 score as our primary optimization metric. While both metrics offer valuable insights, direct comparison should be avoided due to their differing sensitivities to event overlap. A confidence threshold of 0.5 was used to convert model probabilities into binary classifications for both metrics.

**Figure 7:**
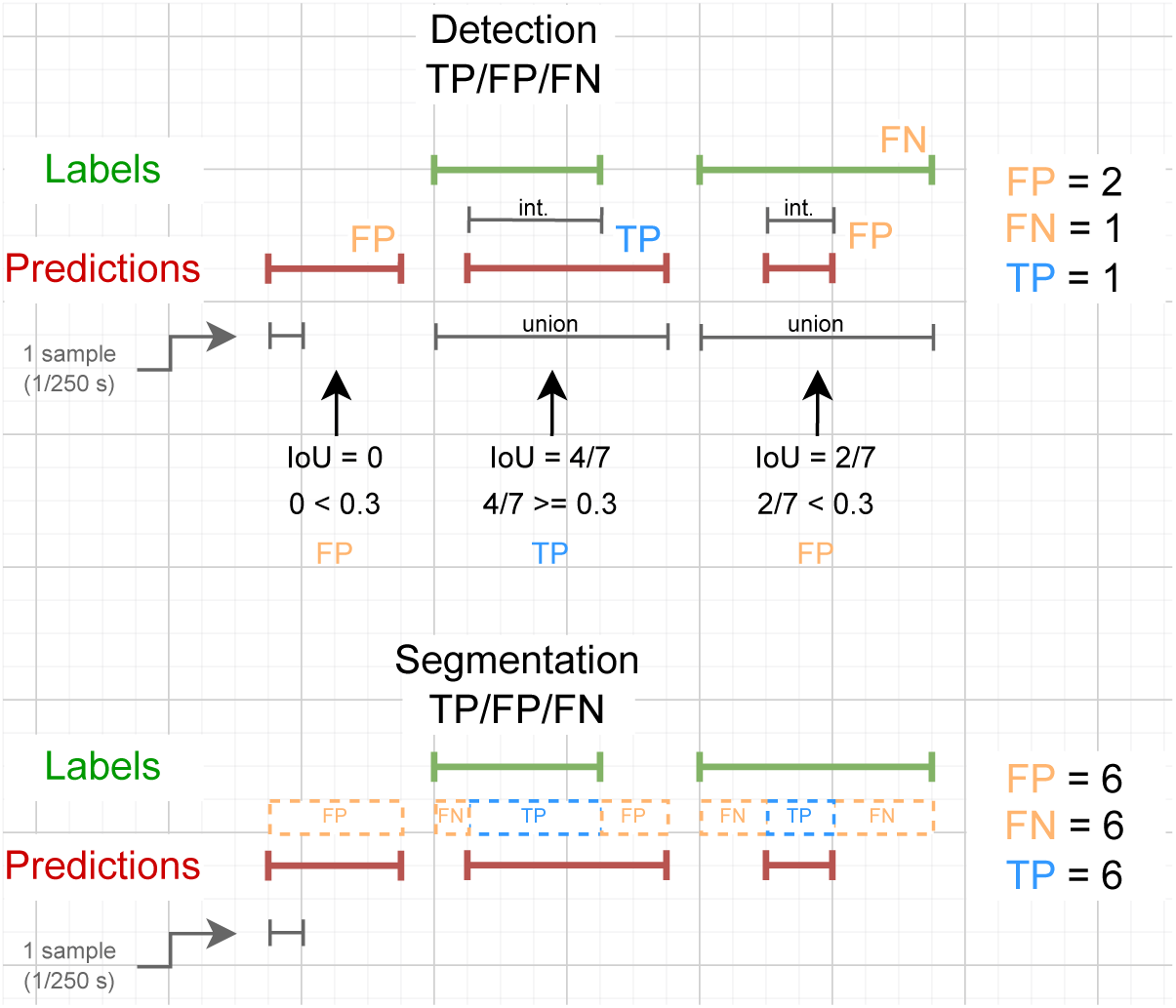
Comparison of the calculations of true positive (TP), false positive (FP), and false negative (FN) counts for computing the detection and segmentation F1 scores. Int. stands for intersection. For clarity, the events shown here are much shorter in duration than real sleep spindles. Notably, in the third prediction, although it is entirely covered by the label (100% segmentation precision), its intersection with the label is too small to be considered a true positive for the detection F1 score.

##### Jaccard Index (JI)

Also known as the Intersection over Union (IoU), this metric measures the overlap between the predicted and ground truth segmentation maps. It is the ratio of the intersection to the union of the predicted and true positive regions, providing another indication of segmentation accuracy. We again set the confidence threshold to 0.5.

##### Average Precision (AP)

Average Precision (AP) summarizes the precision-recall curve by providing a single value representing the area under the curve. This metric is particularly useful for imbalanced data sets and offers insight into the model’s performance at various confidence levels. Unlike the F1 scores and the Jaccard index, which use a specific confidence threshold of 0.5 for the evaluation, AP considers the performance in all possible thresholds, making it a more comprehensive measure of the model’s ability to identify and segment spindles correctly.

##### Area Under the Receiver Operating Characteristic Curve (AUROC)

AUROC measures the model’s ability to differentiate between positive and negative classes across various threshold settings. While AUROC offers a broad view of the model’s performance, we find Average Precision (AP) more relevant for our tasks due to its focus on precision and recall, which are crucial for spindle detection and segmentation due to the events’ scarcity. AUROC can underemphasize the importance of detecting rare positives by treating all threshold ranges equally, which may not reflect the challenges of rare event detection accurately. AP, on the other hand, directly measures performance in detecting rare true positives by focusing on the balance between precision and recall. This makes AP more informative in scenarios like spindle detection among baseline sleep brain waves, where positive cases are rare. By prioritizing AP, we align better with practical applications where the balance between precision and recall is more critical than the overall ranking provided by AUROC.

#### 3.4.2 Segmentation from Detections

Although some of the evaluated models do not explicitly generate a segmentation map for direct evaluation using metrics like the Jaccard index, we can derive a segmentation map from the detections. Specifically, we set detected regions to 1 and all other areas to 0. For the “detection only” variant of our own model, we simulate the segmentation output by averaging the detection confidence values over each of the 30 intervals. In cases where spindle predictions overlap, the segmentation map reflects the average confidence for the corresponding time-steps.

#### 3.4.3 Optimization Techniques

Training the spindle detection model involved several strategies to optimize performance. We used a learning rate finder to dynamically identify the optimal learning rate, which was then adjusted based on the batch size. The batch size was optimized using a batch size tuner, generally set to half the suggested value to avoid overloading GPU memory. Stochastic Weight Averaging (SWA) was utilized to stabilize the parameter shift between epochs and enhance the model’s generalization capabilities, and early stopping with 60-epoch patience was implemented to halt training when validation performance plateaued, thereby reducing the risk of over-fitting. Additionally, we performed hyperparameter optimization (on the validation splits of both datasets) to explore various model configurations and identify the most effective setup. The hyperparameters included the hidden size of the bottleneck representation, dropout rates in the bottleneck and heads, and the head variant itself. We used a PostgreSQL database to store results from Optuna [1] studies, centralizing trial data for accessibility. Optimization was performed through three parallel jobs on MetaCentrum over 24 hours, with results stored in the PostgreSQL database. A tree-structured Parzen Estimator [37] was used to determine the best configurations to explore next, facilitating distributed hyperparameter tuning and accelerating the search for optimal configurations. The training was performed primarily on Nvidia A40 and Tesla T4 GPU cards, with average individual runs taking approximately 25 *±* 6 minutes.

#### 3.4.4 Spindle Characteristics

Beyond mere detection, we developed a tool to extract a set of basic features from each identified sleep spindle, included in our GitHub repository (Section 1). This facilitated a quantitative analysis of spindle morphology, including characteristics such as duration, amplitude, central frequency, density, and spectral power. By aggregating these features across our entire dataset, we were able to derive population-level statistics and assess potential correlations with relevant clinical or demographic variables.

#### 3.4.5 Comparison of Head Variants

After completing the hyperparameter search and selecting the best configuration based on validation performance, we compared the performance of different head variants to determine the optimal architecture for spindle detection and segmentation (see Tables 2a and 2b). This comparison serves as the final step before testing the model on unseen data.

**Table 2:**
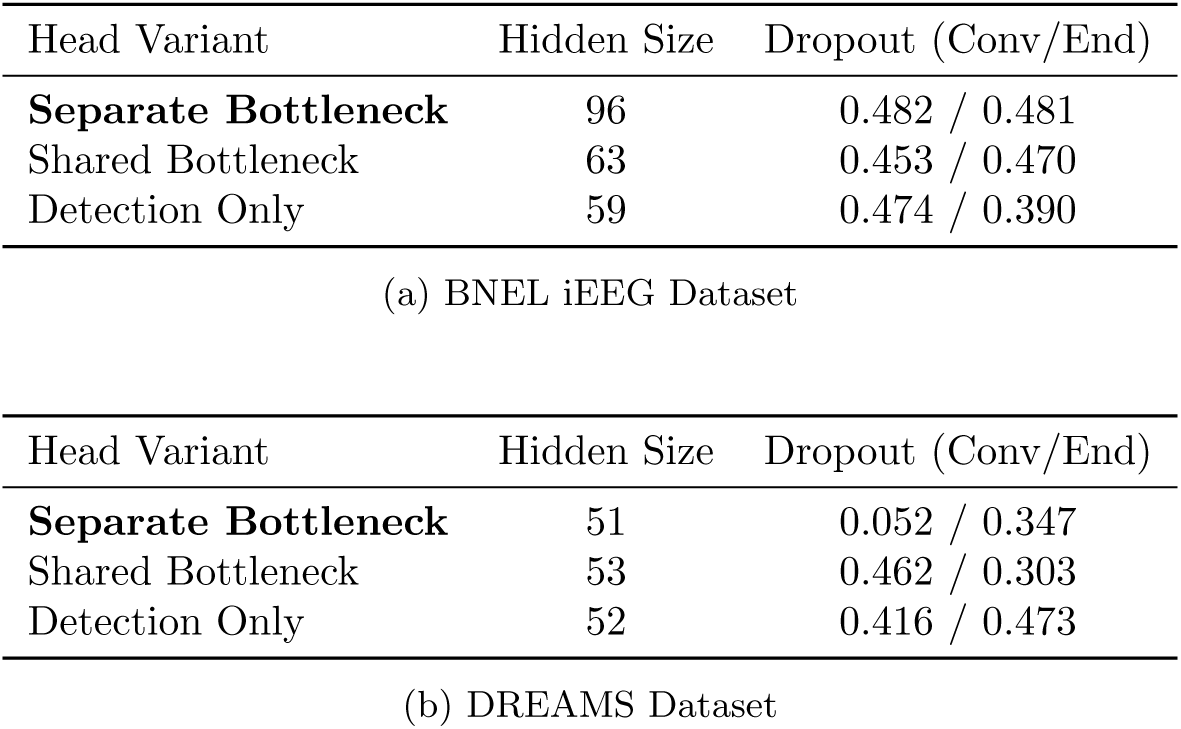
Best hyper-parameter configurations given the validation data. a) for the BNEL iEEG and b) for the DREAMS datasets for each head variant. The separate bottleneck variant, which has shown superior performance, is highlighted.

The comprehensive head-comparison results are presented in Tables A1a and A1b in the Appendix. The following are the key findings.

##### Segmentation

Across both datasets, the dual head variants consistently outperformed the detection-only variant in segmentation. This is reflected in the significant improvements observed in the segmentation F1 score, Jaccard Index (JI) and the average precision (AP) metrics. These results suggest that integrating both detection and segmentation heads enhances the model’s ability to accurately segment spindles. Additionally, the segmentation output can be used to refine the detection boundaries, further improving performance.

##### Detection

For the BNEL iEEG dataset, the separate bottleneck variant demonstrated significant improvements in detection and segmentation AP compared to the other head variants. In particular, it outperformed the detection-only configuration, underscoring its effectiveness in achieving a balanced precision-recall trade-off in detection tasks when trained for both detection and segmentation. The detection-only variant scored better in detection F1 however, which suggests that there is still room for tuning the detection threshold for the separate bottleneck variant.

##### Summary

Our results highlight the advantages of using a dual head approach for spindle oscillation detection and precise segmentation. The results indicate that the separate bottleneck variant can significantly improve both spindle detection and segmentation for specific datasets. Therefore, we have chosen to proceed with the separate bottleneck variant, as it consistently outperformed other methods across multiple metrics.

## 4 Results

Once we identified the best model variant based on validation performance, we proceeded to evaluate it on the test set. The following sections present the outcomes of this evaluation across both datasets.

### 4.1 Evaluation

We evaluated our model’s performance using patient iEEG data, visualized alongside simultaneous scalp EEG recordings in EEG visualization. Figure 8 provides a representative example of our model’s ability to identify sleep spindles in both the anterior nucleus of the thalamus (ANT) and hippocampus (HPC) using intracranial data. This visualization allows for a qualitative assessment of the model’s accuracy, showcasing its ability to detect true spindles while also revealing instances of false negatives (missed spindles) and false positives (incorrectly identified spindles). The simultaneous display of scalp EEG further contextualizes the intracranial activity, demonstrating the model’s capacity to identify spindles that may not be readily apparent in traditional scalp recordings.

**Figure 8:**
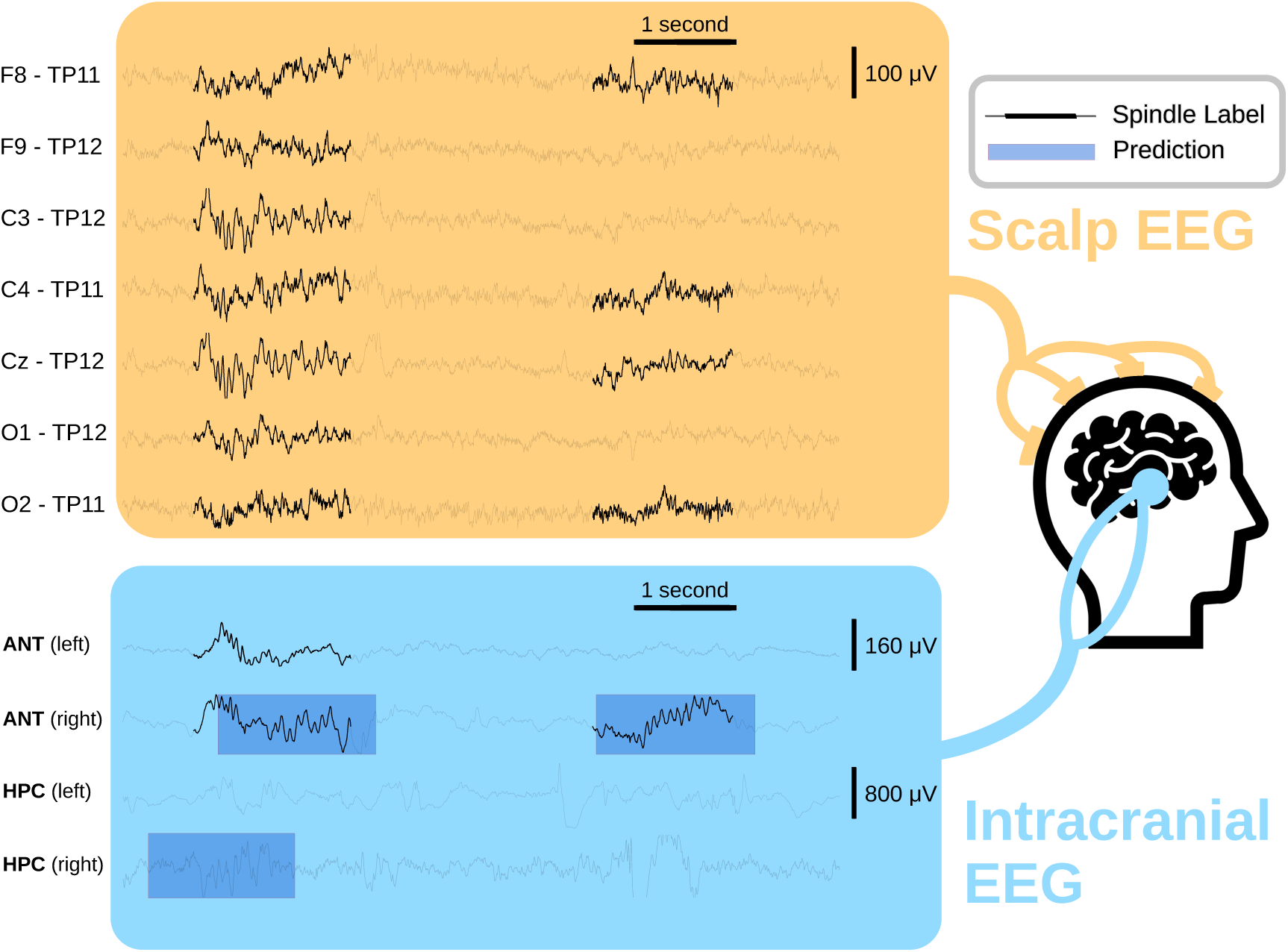
Model inference on patient iEEG data. Intracranial EEG (iEEG) recordings are shown alongside synchronously recorded scalp EEG. Spindles (*Spindle Label*) were marked by an expert in both. Several spindles span multiple channels simultaneously, including across iEEG and scalp EEG. The model, however, was applied only to iEEG data. Blue *Prediction* markers indicate spindle detections (confidence *>* 0.5, IoU *<* 0.3) in the anterior nucleus of the thalamus (ANT) and hippocampus (HPC). The figure highlights a false positive (right HPC—prediction without label) and a false negative (left ANT—label without prediction). Interestingly, the false positive in the right HPC aligns with manually labeled events in both the scalp EEG and right ANT, suggesting possible spindle activity that was not explicitly annotated in the HPC.

We evaluated the performance of our proposed method by comparing it to several state-of-the-art sleep spindle detection algorithms. Of the methods reviewed in Section 2, YASA, A7, and SUMO were publicly available and implemented in Python, enabling direct comparison on our dataset. While other published approaches [7, 30, 23, 38] were evaluated on the DREAMS dataset, we leveraged this publicly available dataset as an additional benchmark. This allowed us to assess our method’s performance against these previously reported results, providing a broader context for evaluating its effectiveness.

Our model performs robustly on both the iEEG and DREAMS datasets, as shown in Figure 9, where the receiver operating characteristic (ROC) and precision-recall (PR) curves illustrate its effectiveness in accurately identifying and delineating sleep spindles. Detailed analyses of these results, alongside comparisons to other approaches in both datasets, are presented in the following sections.

**Figure 9:**
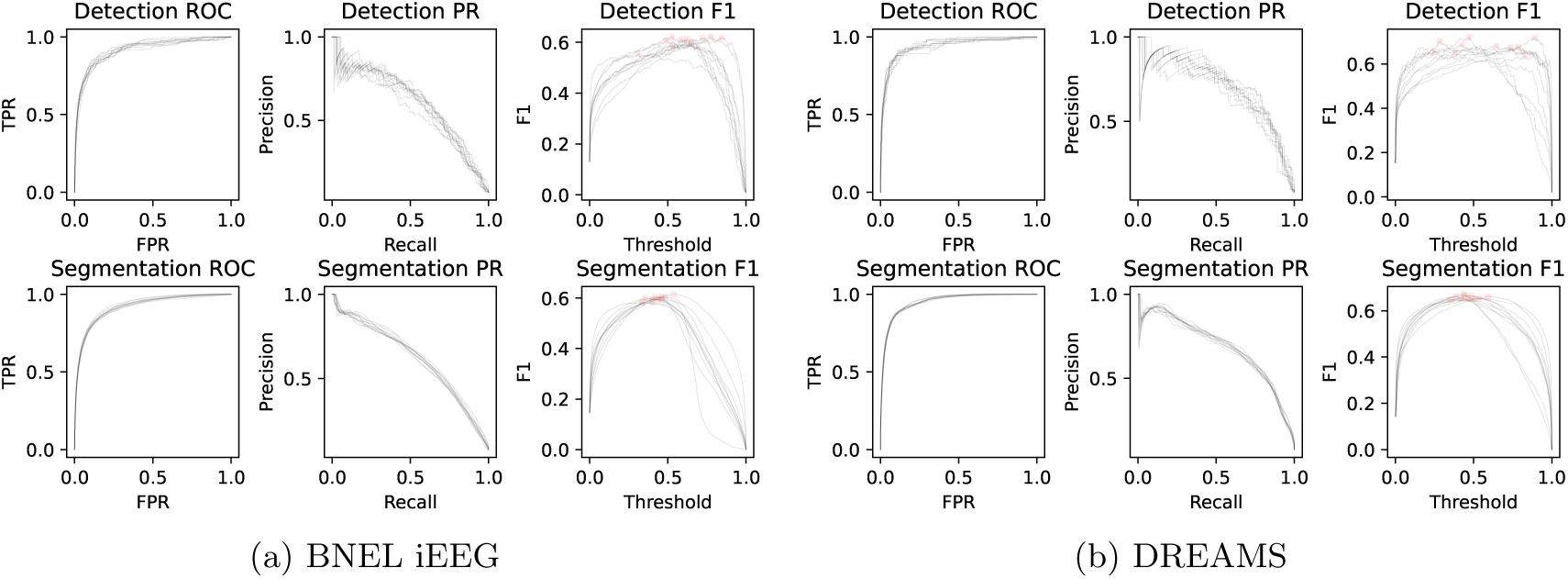
Receiver operating characteristic (ROC), precision-recall (PR), and F1 threshold curves. ROC curves show the trade-off between true positive rate (sensitivity) and false positive rate across different decision thresholds. PR curves illustrate the relationship between precision (positive predictive value) and recall (sensitivity), which is particularly informative for imbalanced datasets. F1 threshold curves represent the F1 score — the harmonic mean of precision and recall — calculated at varying decision thresholds between 0 and 1. The curves correspond to our best model evaluated on (a) the intracranial BNEL EEG dataset on the left-hand side and on (b) the DREAMS scalp EEG dataset on the right-hand side. For metric details, see Section 3.4.1.

#### 4.1.1 BNEL iEEG

Table 3 summarizes the performance of various methods we evaluated on the BNEL iEEG test set. Our method achieves the highest segmentation F1 score (**0.59** *±* 0.01) and further excels in detection F1 (**0.67** *±* 0.02) and both average precision metrics (**0.62** *±* 0.01 for segmentation, **0.60** *±* 0.01 for detection), demonstrating its overall solid performance in detection and segmentation of intracranial sleep spindles. An example of our model predictions on multi-channel iEEG data is visualized in Figure 8, and an example of isolated and detailed predictions on single-channel iEEG is visualized in Figure 4a.

**Table 3:**
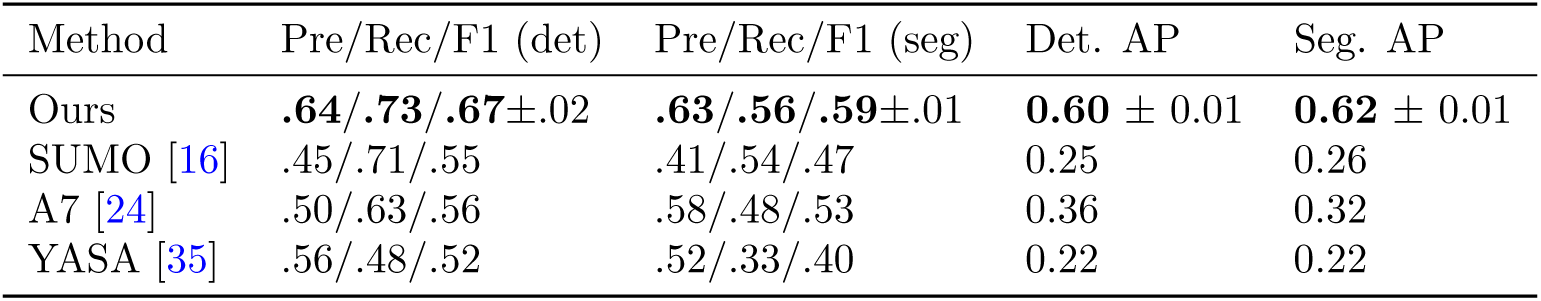
Results on intracranial BNEL iEEG dataset. The highest values are marked in **bold** text. Values corresponding to stochastic methods include the 95% confidence interval, except for precision and recall values of our method, which are provided separately in Table A2 for clarity. The evaluated metrics, commonly used in the literature, are described in Section 3.4.1.

Other methods, such as SUMO and A7, show competitive precision and recall values, as shown in Table 3, but their F1 scores fall short compared to ours. For example, SUMO achieves similar recall (0.71 vs. 0.73 in detection, 0.54 vs. 0.56 in segmentation), yet its F1 score is lower than ours (0.55 vs. 0.67 in detection, 0.47 vs. 0.59 in segmentation) indicating a trade-off between precision and recall; specifically, SUMO marks many non-spindles as spindles, causing higher rates of false positives. The YASA method also shows areas of strength, but does not surpass our method in any of the key metrics.

#### 4.1.2 DREAMS

Tables 4 and 5 present the results on the DREAMS dataset. Our method achieves the highest detection F1 score (**0.69** *±* 0.02), demonstrating a balanced trade-off between precision and recall. While DETOKS and McSleep report slightly higher segmentation F1 scores (**0.70** *±* 0.02 and 0.66 *±* 0.02, respectively), their performance is dependent on manually and heuristically tuned threshold-like parameters. In contrast, our approach is fully automatic, with all parameters selected strictly on non-test data. Notably, DETOKS and McSleep rely on empirically chosen thresholds that appear to have been adjusted with knowledge of the test set, raising concerns about their generalizability. Since our method avoids manual tuning entirely, it is more robust and better suited for real-world deployment across different datasets.

**Table 4:** Results on the scalp EEG DREAMS dataset. Best values are in **bold** text. Values corresponding to stochastic methods include the 95% confidence interval, except for precision and recall values of our method, which are provided separately in Tables 5 and A3 for clarity. The evaluated metrics, commonly used in the literature, are described in Section 3.4.1.

| Method | Pre/Rec/F1 (det) | Pre/Rec/F1 (seg) | Det. AP | Seg. AP |
| --- | --- | --- | --- | --- |
| Ours | .68/**.73/.69**±.02 | .65/**.65/.65** ± 0.01 | **0.70** ± 0.01 | **0.68** ± 0.01 |
| SUMO [16] | .72/.45/.55 | .63/.31/.42 | 0.37 | 0.25 |
| A7 [24] | .71/.52/.60 | .70/.41/.52 | 0.35 | 0.33 |
| YASA [35] | **.82**/.15/.26 | **.76**/.12/.21 | 0.18 | 0.16 |

**Table 5:**
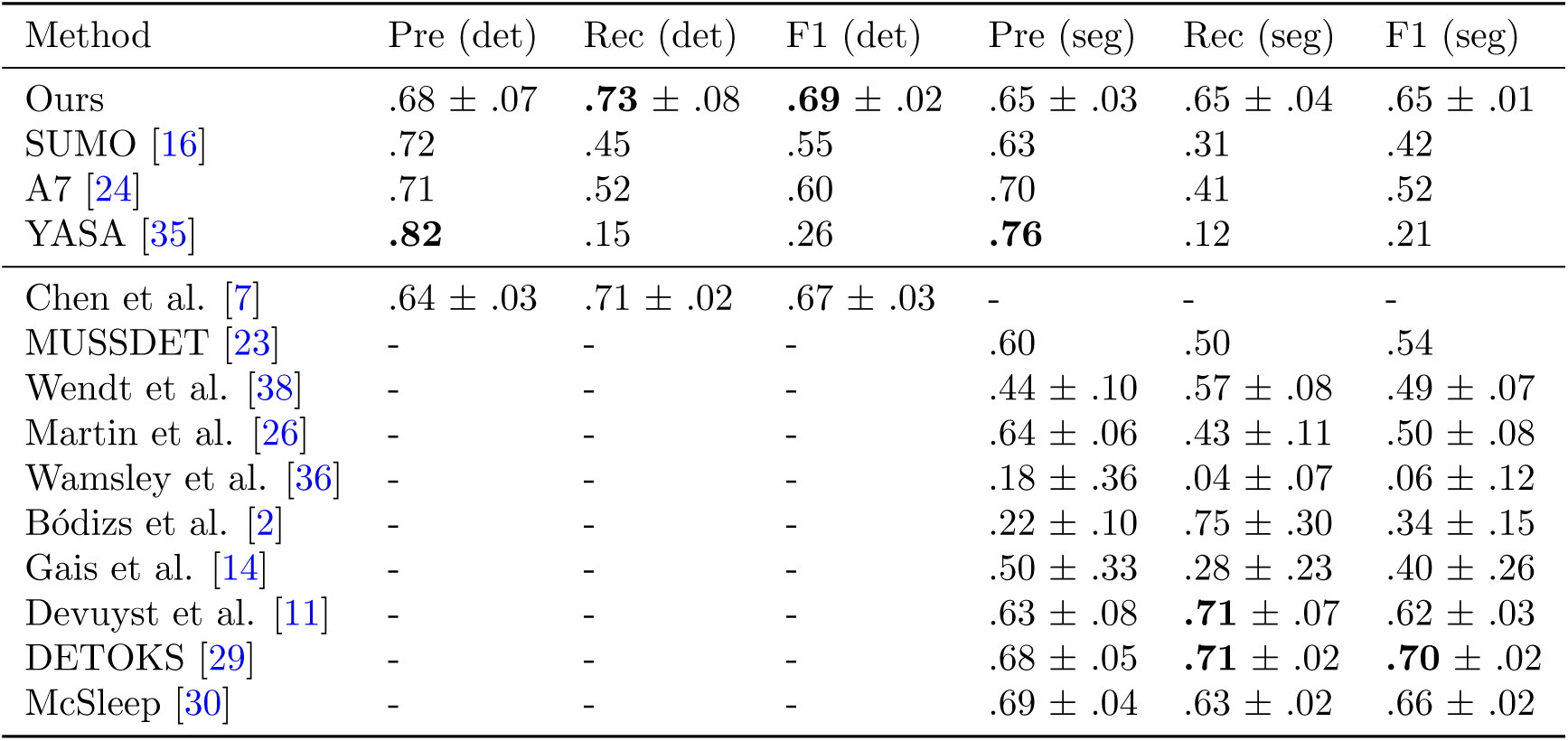
Precision, recall, and F1 scores for the scalp EEG DREAMS dataset. The upper part was calculated by us, the bottom part is as reported by Chen et al. [7] and Parekh et al. [30]. Best values are in **bold** text. Values corresponding to stochastic methods include the 95% confidence interval. Metrics are described in Section 3.4.1.

Among the other methods, YASA stands out with an exceptionally high detection precision (**0.82**) but a strikingly low recall (0.15), indicating that it applies an extremely strict threshold to avoid false positives but consequently misses the majority of actual events. This suggests that YASA may be useful in settings where high confidence is prioritized over completeness but is poorly suited for comprehensive detection. Similarly, SUMO and A7 show relatively strong precision but much lower recall, leading to detection F1 scores of 0.55 and 0.60, respectively, significantly lower than our approach.

For segmentation, Devuyst et al. achieve the highest recall (**0.71**), but their precision (0.63) is lower than that of DETOKS and McSleep, which both maintain a better balance between precision and recall. Some classical methods, such as those by Wamsley et al. and Bódizs et al., exhibit highly imbalanced performance, with either extremely low recall (Wamsley et al.: 0.04) or extremely low precision (Bódizs et al.: 0.22), making them unsuitable for practical use.

In summary, our proposed method demonstrates superior performance in both scalp and intracranial EEG datasets, achieving high accuracy in both spindle detection and segmentation. Unlike existing methods that either prioritize precision at the cost of recall (e.g., YASA) or rely on manually tuned parameters (e.g., DETOKS, McSleep), our approach maintains a consistently high balance across both metrics without requiring manual intervention. This fully automatic framework ensures greater generalizability, robustness, and adaptability across different datasets. Notably, our model not only matches existing state-of-the-art methods in scalp EEG spindle analysis but also enables automated and reliable spindle detection in intracranial EEG recordings, opening new avenues for investigating sleep spindles in both research and clinical settings.

### 4.2 Sleep Spindle Characteristics

Table 6b summarizes the spectral and temporal characteristics of sleep spindles computed for the ground truth labels and spindles predicted by our model in both datasets used in our study. These characteristics provide insight into the general properties of these datasets and the quality of predicted spindles with respect to the annotated ground truth in terms of these signal properties.

**Table 6:**
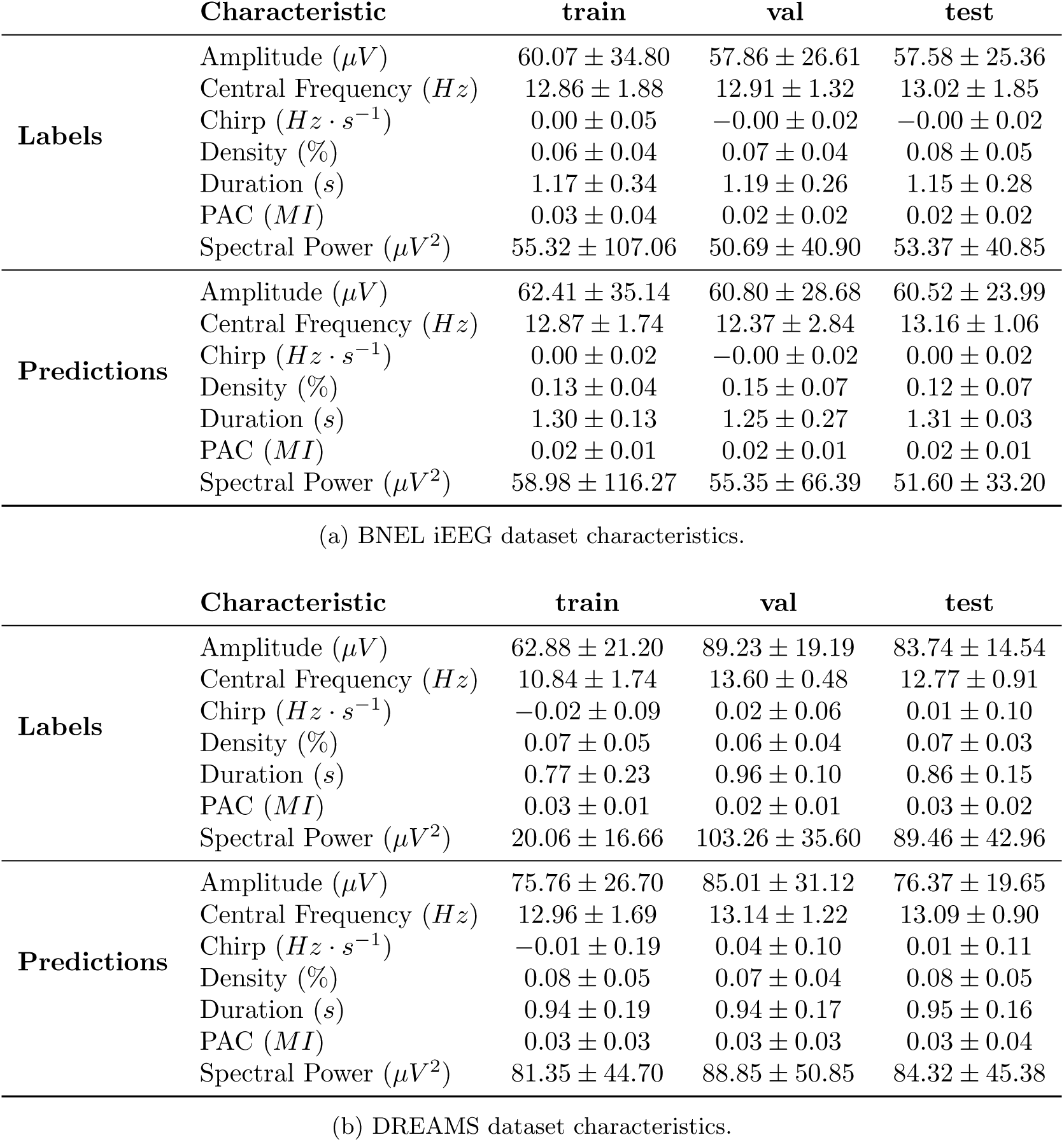
Characteristics of sleep spindles annotated by human annotators (*Labels*) compared to those predicted by our model (*Predictions*, best given validation data). The displayed values are formatted as *mean ± std*. Density percentage stands for the amount of signal covered by spindles across all 30-second intervals in the dataset. PAC stands for (sleep spindle-slow wave) Phase-Amplitude Coupling.

#### Amplitude

For the BNEL iEEG dataset, the amplitudes of predicted spindle events closely approximate ground truth across all splits, with minor deviations (57.58 *±* 25.36 *µV* vs. 60.52 *±* 23.99 *µV*). This indicates consistent detection of events with similar amplitude spans. For the DREAMS dataset, the predicted spindle intervals have slightly lower amplitudes compared to the ground truth in the test set (83.74*±*14.54 *µV* vs. 76.37*±*19.65 *µV*). This may reflect missed high-amplitude spindles or imprecise boundaries that underestimate the peak signal amplitudes. Longer spindles can reduce the average spindle amplitude, as the total signal power is spread over a longer interval. However, this effect should not impact the central amplitude, which corresponds to the peak of the spindle where the signal is expected to be maximal. If the model preserves central amplitude accuracy while predicting slightly lower average amplitudes, this could suggest that the predicted spindle boundaries are slightly extended rather than a true underestimation of peak signal strength. Further analysis of spindle duration and intra-spindle amplitude variation could clarify this distinction.

#### Central Frequency

For the BNEL iEEG dataset, central frequencies of predicted spindles show strong agreement with the ground truth across all splits, even in the test set (13.02 *±* 1.85 *Hz* vs. 13.16*±*1.06 *Hz*). In the DREAMS dataset, predictions demonstrate close alignment with labels in the validation (13.14*±*1.22 *Hz* vs. 13.60*±*0.48 *Hz*) and test sets (13.09*±*0.90 *Hz* vs. 12.77 *±* 0.91 *Hz*). However, the train set reveals a systematic discrepancy, with predictions exhibiting higher frequencies than labels (12.96 *±* 1.69 *Hz* vs. 10.84 *±* 1.74 *Hz*). This contrast suggests that while the model generally captures the dominant frequency range of sleep spindles, it appears to overestimate central frequencies in training data containing lower-frequency spindle examples. This could also suggest a minor over-fit to the validation data frequency spectrum, as the model was early-stopped on this particular split. The preserved alignment in validation and test sets indicates robustness to frequency variations within the target 9-18 Hz range.

It is worth noting that spindle research generally distinguishes between slow (*<* 13 Hz) and fast (*≥* 13 Hz) spindles. If the observed discrepancy does not significantly alter the proportion of slow versus fast spindles, the overestimation in central frequency during training may have limited practical impact on spindle detection. Further analysis of the predicted spindle frequency distribution could clarify whether the model maintains accurate classification between slow and fast spindles despite minor frequency deviations.

#### Density

In the BNEL iEEG dataset, the predicted spindle density is slightly higher on average than the ground truth across all splits, particularly in the training set (0.06 *±* 0.04% vs. 0.13 *±* 0.04%), suggesting that increasing the detection confidence threshold could improve precision in deployment. This is further confirmed in Tables 3 and A2, where recall is significantly higher than precision for our method. A similar but less pronounced pattern is present in the DREAMS dataset, where the model more accurately captures spindle density in scalp recordings. The difference in spindle density between predictions and ground truth in the BNEL iEEG data likely results from factors such as missed detections by human annotators and the presence of artifacts in the iEEG recordings that share spectral similarities with sleep spindles.

#### Duration

In both datasets, predicted spindle duration is slightly longer compared to the ground truth events (1.15 *±* 0.28 *s* vs. 1.31 *±* 0.03 *s* for the BNEL iEEG test set, 0.86 *±* 0.15 *s* vs. 0.95 *±* 0.16 *s* for the DREAMS test set). This suggests that the model may struggle to correctly adapt to very short spindles, potentially due to architectural limitations. A more flexible head design could allow better adaptation by providing greater freedom in selecting the start and end points of detected intervals.

#### Spectral Power

Across both datasets and all splits, the predicted spectral power values closely align with the ground truth, with minor deviations observed. In the BNEL iEEG test set, the predicted spectral power is 51.60 *±* 33.20*, µV* ^2^, compared to the ground truth of 53.37 *±* 40.85*, µV* ^2^. Similarly, in the DREAMS test set, predictions yield 84.32 *±* 45.38*, µV* ^2^ against a ground truth of 89.46 *±* 42.96*, µV* ^2^. These results indicate that the model effectively captures the spectral characteristics of sleep spindles in both intracranial and scalp EEG recordings. The slight underestimation of spectral power in the DREAMS dataset suggests a conservative bias in detecting spindle amplitudes in scalp EEG, while the near-equivalence in the BNEL iEEG data reflects the model’s ability to generalize well across different recording modalities.

Overall, the model demonstrates robust performance across multiple spindle characteristics in both the BNEL iEEG and DREAMS datasets. Predicted spindle amplitudes and spectral power closely approximate ground truth values, with minor underestimations in the DREAMS dataset, likely reflecting conservative detection in scalp EEG. Central frequencies align well across datasets, indicating the model’s proficiency in capturing dominant spindle frequencies. While spindle densities are slightly overestimated in BNEL iEEG, particularly in training data, this may stem from artifacts or annotation inconsistencies, and adjusting detection thresholds could improve precision. Finally, predicted spindle durations tend to be longer than ground truth, indicating potential model limitations in capturing shorter events, which might be addressed through architectural adjustments.

### 4.3 Limitations

While our approach shows promising results on the DREAMS and BNEL iEEG datasets, it is not without its limitations. As depicted in Figures 4 and 8, the model sometimes generates false negatives and false positives. False negative cases generally arise when spindles are too faint to noticeably alter the spectral properties of the signal, making them challenging to distinguish from the baseline EEG. False positives occur when artifacts or noise in the EEG signal display spectral characteristics similar to spindles or similarly affect the scalogram, as clearly seen in Figure 4. These drawbacks emphasize the difficulties in differentiating spindles from short bursts of noise or artifacts that have overlapping frequency traits. The longer noisy signals are differentiated well due to their extended temporal structure, which allows the model to recognize inconsistent spectral patterns over time – a feature not present in short-duration spindles (typically 0.5-3 seconds). However, this duration-dependent discrimination leaves the model vulnerable to brief (*<* 1 second) artifacts that spectrally resemble spindles, particularly in channels with inherent physiological noise (e.g., muscle activity or electrode movement). Future work could mitigate these issues by explicitly modeling temporal stability constraints or incorporating multi-scale context through hierarchical architectures.

Additionally, the analysis of spindle characteristics reveals key areas for refinement. Although the model demonstrates strong results on the scalp-specific DREAMS dataset, achieving good alignment with the ground truth does not fully resolve the challenges unique to that dataset. The DREAMS dataset, characterized by a relatively stable signal-to-noise ratio and consistent spindle annotation criteria, allows for better alignment in terms of evaluation metrics and spectral properties. However, this raises concerns about the model’s applicability to other datasets or real-world scenarios, where EEG data might exhibit greater noise and spectral variations. Specifically, the model’s tendency to predict higher spindle amplitudes and spectral power than the ground truth suggests potential issues in detecting low-amplitude spindles or defining accurate intervals, which may not be as prevalent in the controlled environment of the DREAMS dataset.

Note that detecting spindles in thalamic leads may be different than detecting spindles in other intracranial leads away from the thalamus. Spindle generation is closely linked to thalamocortical circuits, and spindles recorded directly from thalamic leads may differ in amplitude, duration, and spectral characteristics compared to those detected in cortical or other intracranial leads. This difference could further complicate generalization across datasets and recording modalities.

Furthermore, the model’s slight overestimation of spindle duration and the higher density of predicted events in the BNEL iEEG dataset indicate areas for improvement, particularly in adapting to very short spindles and addressing false positives. These findings suggest that while the model performs well in controlled conditions, additional work is needed to improve precision and adaptability in noisier, more variable datasets. Future enhancements could focus on:

- **Refining boundary detection** to improve the accuracy of spindle duration, amplitude, and coupling, especially in challenging cases with faint or brief spindles.
- **Improving artifact filtering** to minimize false positives caused by noise and other non-spindle signals, ensuring that only true spindles are identified.
- **Expanding the diversity of training datasets** to better capture the variability of EEG signals across different conditions and populations, which would improve the model’s robustness in real-world settings.

These efforts could significantly improve the robustness and generalizability of spindle detection performance, paving the way for broader applications in clinical and research settings.

## 5 Conclusion

In this work, we present a novel approach for detecting and segmenting sleep spindles using established machine learning techniques. We compared our method to various state-of-the-art methods on multiple datasets. While we were primarily focusing on iEEG data, our method demonstrates strong performance across two diverse scenarios, iEEG and scalp EEG sleep data. In the iEEG data, our approach achieves the highest F1 score and excels in spindle detection and segmentation average precision, highlighting its ability to effectively balance precision and recall.

Our approach provides a reliable and effective solution for spindle detection and segmentation, outperforming existing methods in key performance metrics. The technique can open new avenues and directions in research to improve our understanding of brain functions and health and pave the way to techniques that can be used in clinical settings to improve patient care.

## CRediT author statement

**Michal Sejak**: Conceptualization, Methodology, Software, Validation, Formal analysis, Investigation, Data curation, Visualization, Writing – original draft. **Filip Mivalt**: Conceptualization, Software, Resources, Visualization, Writing – review & editing. **Vladimir Sladky**: Writing – review & editing. **Vit Vsiansky**: Data curation, Investigation. **Diego Z. Carvalho**: Writing – review & editing. **Erik K. St. Louis**: Writing – review & editing. **Gregory A. Worrell**: Writing – review & editing. **Vaclav Kremen**: Conceptualization, Supervision, Resources, Writing – original draft, Writing – review & editing.

## Data Availability

We evaluated our spindle detection algorithm on two datasets: the publicly available DREAMS
database (https://zenodo.org/records/2650142) and the proprietary Bioelectronics Neurophysiology and Engineering Lab (BNEL)
intracranial EEG (iEEG) dataset. The BNEL iEEG dataset was collected at the Mayo Clinic
and is available from the authors upon reasonable request. Additional details about the BNEL lab can be found at
https://www.mayo.edu/research/labs/bioelectronics-neurophysiology-engineering/overview.

https://github.com/msel-source/pymef

https://best-toolbox.readthedocs.io/en/latest/index.html

## Acknowledgments

The authors thank people with epilepsy for participating in this research. The authors thank Certicon a.s. for the use of the CyberPSG tool for the visual review of EEG. The authors thank Pavla Havánová for her valuable feedback on the design and clarity of the diagrams. Computational resources were provided by the e-INFRA CZ project (ID:90254), supported by the Ministry of Education, Youth and Sports of the Czech Republic.

This work has been supported by the Grant no. SGS-2025-022 - New Data Processing Methods in Current Areas of Computer Science.

---

**A Optimal Variant Selection**

**B Detailed Results**

**Table A1:**
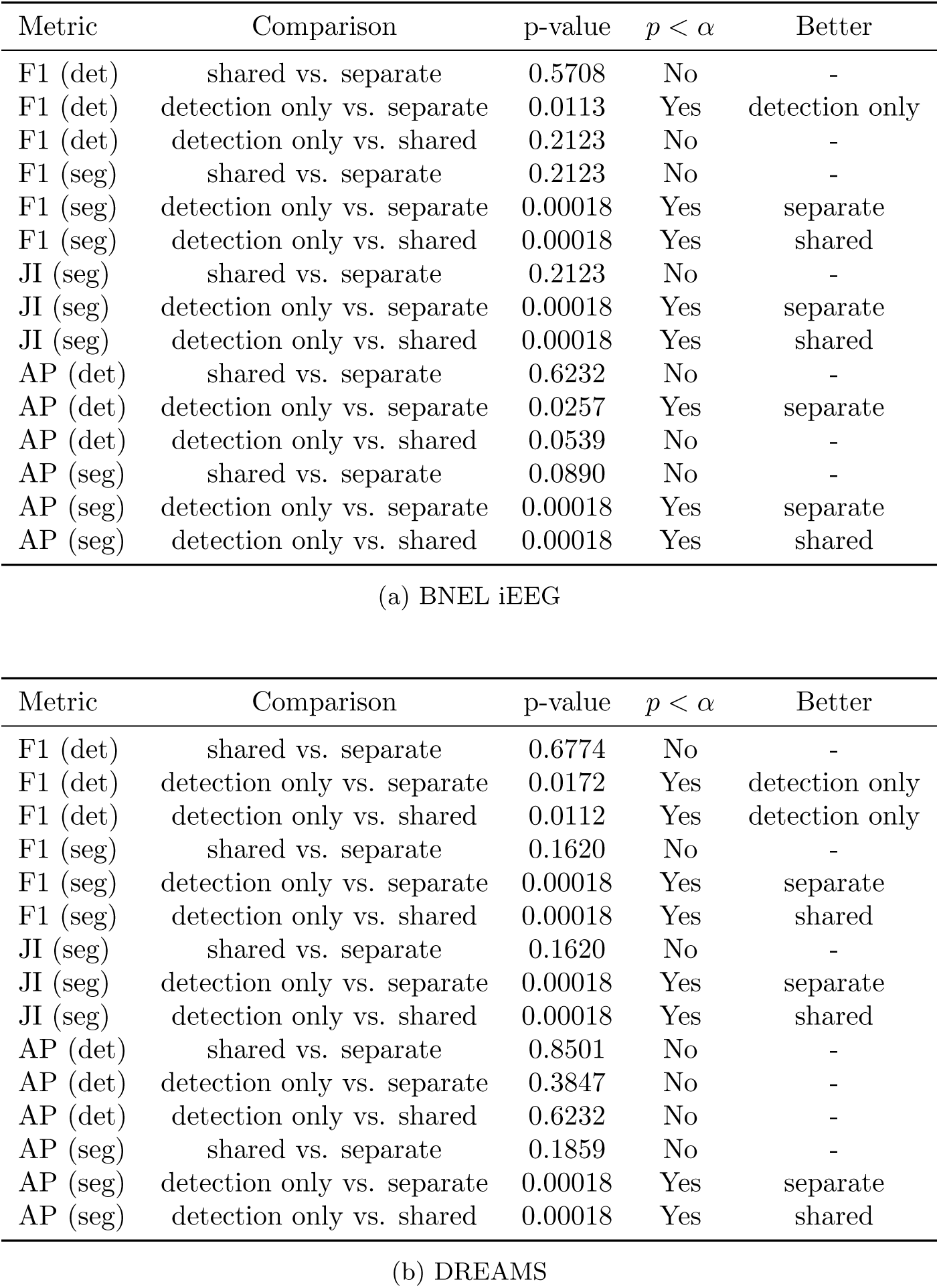
Detailed head variant evaluation results. We used the two-sided Mann-Whitney U test for determining whether one variant of the model head outperforms the other. We chose *α* = 0.05 as our level of significance. See Section 3.4.1 for metric descriptions and Section 3.3.3 for head variant descriptions.

**Table A2:** Results on BNEL iEEG dataset. Best values are bolded. Values corresponding to stochastic methods include the 95% confidence interval.

| Metric | Ours | Sumo [16] | A7 [24] | YASA [35] |
| --- | --- | --- | --- | --- |
| Pre (det) | **0.64 (0.04)** | 0.45 (-) | 0.50 (-) | 0.56 (-) |
| Rec (det) | **0.73 (0.05)** | 0.71 (-) | 0.63 (-) | 0.48 (-) |
| F1 (det) | **0.67 (0.02)** | 0.55 (-) | 0.56 (-) | 0.52 (-) |
| Pre (seg) | **0.63 (0.03)** | 0.41 (-) | 0.58 (-) | 0.52 (-) |
| Rec (seg) | **0.56 (0.04)** | 0.54 (-) | 0.48 (-) | 0.33 (-) |
| F1 (seg) | **0.59 (0.01)** | 0.47 (-) | 0.53 (-) | 0.40 (-) |
| JI (seg) | **0.41 (0.01)** | 0.31 (-) | 0.36 (-) | 0.25 (-) |
| AUROC (det) | **0.92 (0.01)** | 0.76 (-) | 0.77 (-) | 0.67 (-) |
| AP (det) | **0.60 (0.01)** | 0.25 (-) | 0.31 (-) | 0.22 (-) |
| AUROC (seg) | **0.92 (0.00)** | 0.74 (-) | 0.73 (-) | 0.65 (-) |
| AP (seg) | **0.62 (0.01)** | 0.26 (-) | 0.32 (-) | 0.22 (-) |

**Table A3:** Results on DREAMS dataset. Best values are bolded. Values corresponding to stochastic methods include the 95% confidence interval.

| Metric | Ours | Sumo [16] | A7 [24] | YASA [35] |
| --- | --- | --- | --- | --- |
| Pre (det) | 0.68 (0.07) | 0.72 (-) | 0.71 (-) | **0.82 (-)** |
| Rec (det) | **0.73 (0.08)** | 0.45 (-) | 0.52 (-) | 0.15 (-) |
| F1 (det) | **0.69 (0.02)** | 0.55 (-) | 0.60 (-) | 0.26 (-) |
| Pre (seg) | 0.65 (0.03) | 0.63 (-) | 0.70 (-) | **0.76 (-)** |
| Rec (seg) | **0.65 (0.04)** | 0.31 (-) | 0.41 (-) | 0.12 (-) |
| F1 (seg) | **0.65 (0.01)** | 0.42 (-) | 0.52 (-) | 0.21 (-) |
| JI (seg) | **0.48 (0.01)** | 0.27 (-) | 0.35 (-) | 0.12 (-) |
| AUROC (det) | **0.94 (0.01)** | 0.71 (-) | 0.73 (-) | 0.57 (-) |
| AP (det) | **0.70 (0.01)** | 0.37 (-) | 0.35 (-) | 0.18 (-) |
| AUROC (seg) | **0.95 (0.00)** | 0.65 (-) | 0.70 (-) | 0.56 (-) |
| AP (seg) | **0.68 (0.01)** | 0.25 (-) | 0.33 (-) | 0.16 (-) |

